# Immunity response against mild-to-moderate breakthrough COVID-19

**DOI:** 10.1101/2022.05.30.22275050

**Authors:** Pichanun Mongkolsucharitkul, Apinya Surawit, Nitat Sookrung, Anchalee Tungtrongchitr, Pochamana Phisalprapa, Naruemit Sayabovorn, Weerachai Srivanichakorn, Chaiwat Washirasaksiri, Chonticha Auesomwang, Tullaya Sitasuwan, Thanet Chaisathaphol, Rungsima Tinmanee, Methee Chayakulkeeree, Pakpoom Phoompoung, Watip Tangjittipokin, Sansanee Senawong, Gornmigar Sanpawitayakul, Saipin Muangman, Korapat Mayurasakorn, the SPHERE Investigators

**Affiliations:** Siriraj Population Health and Nutrition Research Group, Department of Research Group and Research Network, Faculty of Medicine Siriraj Hospital, Mahidol University; Center of Research Excellence on Therapeutic Proteins and Antibody Engineering, Department of Parasitology, Faculty of Medicine Siriraj Hospital, Mahidol University; Division of Ambulatory Medicine, Department of Medicine, Faculty of Medicine Siriraj Hospital, Mahidol University; Division of Infectious Diseases and Tropical Medicine, Department of Medicine, Faculty of Medicine Siriraj Hospital, Mahidol University; Department of Immunology, Faculty of Medicine Siriraj Hospital, Mahidol University; Division of Ambulatory Paediatrics, Department of Paediatrics, Faculty of Medicine Siriraj Hospital, Mahidol University; Department of Anesthesiology, Faculty of Medicine Siriraj Hospital, Mahidol University

## Abstract

**BACKGROUND:** The Omicron variant prevails the Delta variant after December 2021 in Thailand. Both variants of concern embody diverse epidemiological trends and immunogenicity, raising enormous public health concerns. We determined whether biological and clinical characteristics and immunogenicity of patients differ between Delta and Omicron during post-coronavirus disease 2019 (COVID-19) stage.

**METHODS:** A retrospective cohort study involved patients with mild-to-moderate COVID-19 who were under a home isolation (HI) strategy. Clinical outcomes and laboratory data of 2704 and 2477 patients during the Delta and Omicron pandemics were analyzed, respectively. We evaluated anti-receptor binding domain immunoglobulin G (anti-RBD IgG) and surrogate viral neutralizing (sVNT) activity in a subset of 495 individuals post-COVID-19 infection during the Delta pandemic.

**RESULTS:** Eighty-four percent of all patients received antiviral treatment. The peak cycle threshold (Ct) values, which inversely related to viral load, were lower in the Omicron (19 [IQR=17-22]) compared with the Delta (21 [IQR=18-26]; p<0.001), regardless of vaccination status. Upper respiratory tract symptoms were common signs during the Omicron compared with the Delta pandemic. At least two-dose vaccination reduced the chance of hospital readmissions by 10–30% and death by less than 1%. Furthermore, anti-RBD IgG and sVNT against the Delta variants tended to be higher among the older individuals after post-COVID 19 infections and expressed in the long interval after two-dose vaccination than in other groups.

**CONCLUSIONS:** Mild-to-moderate Delta and Omicron breakthrough infection with prior full vaccination is limitedly immunogenic; thereby exerting reduced protection against reinfection and infection from novel variants. However, this may be only sufficient to prevent hospitalization and death, particularly in countries where vaccines are limited. (ClinicalTrials.gov number, NCT05328479.)

## INTRODUCTION

Thailand faced rapidly third and fourth waves of coronavirus disease 2019 (COVID-19) outbreak since 2021. Social distancing and vaccines are encouraged as a survival tool for people to circumvent this threat.^1,2^ CoronaVac (Sinovac, Life Science) and ChAdOx1 (AstraZeneca/Oxford) were widely used than other vaccines for most Thais with limited supply since February 2021.^3^ Both vaccines were efficacious against symptomatic COVID-19 caused by the Wuhan strain but lower extent against other variants of concern including Delta and Omicron variants.^4,5^ Currently, the Omicron variant displaced the Delta variant during the study period.^1^ Random selection for severe acute respiratory syndrome coronavirus 2 (SARS-CoV-2) variants surveillance from Department of Medical Science^4^ and worldwide during the week 4-10 of 2022 demonstrated that almost all new infections in Thailand are from the Omicron variant (99.6%).

Combination of waning vaccine-derived immunity and an arrival of SARS-CoV-2 variant: the Delta variant (B.1.617.2) and the Omicron variant (B.1.1.529), led to breakthrough infections after COVID-19 vaccination or prior infection.^5,6^ This greatly overloaded the nation’s public health system and exacerbated socioeconomic disparities.^2,7^ In response, home isolation (HI) strategy has been implemented for patients with mild to moderate symptoms nationwide to combat an overwhelming demand for hospital beds (Method S1).

Serious concerns about the Omicron variant have been drawn due to its increased transmissibility^8^ and a potential for reduced sensitivity to neutralizing antibodies associated with immune escape, and new emerging Omicron lineages (BA.4 and BA.5),^9-11^ although Omicron causes less severe disease than other variants.^12^ Little are data available on an early presentation in patients with mild-to-moderate severity and post-infection immunogenicity of patients, and very few studies verify the guidance for vaccination after mildly-moderately COVID-19 infection particularly in countries where fully vaccination is challenging tasks.^13^ We demonstrated here completed results of retrospective information of mild-moderate symptomatic individuals who were seropositive for SARS-CoV-2 and were treated in HI system from July 2021 to March 2022 during the Delta and Omicron pandemics.

## METHODS

### STUDY DESIGN AND POPULATION

A retrospective cohort study was conducted to evaluate treatment outcomes and immunogenicity of mild to moderate COVID-19 among patients who were admitted in HI system of Siriraj Hospital, the largest university hospital in Bangkok, Thailand (details were provided in the Supplementary Appendix). The data collection reported here was performed between July 8, 2021 to March 15, 2022. The study population included 2704 and 2477 patients during the Delta (before November 2021) and Omicron (after January 12, 2022) pandemics who had been positive with SARS-CoV-2 as determined by a reverse transcriptase polymerase chain reaction (RT-PCR) testing. Data were retrieved from electronic medical record of each patient, which included demographic, clinical information, clinical manifestations, and laboratory test findings. A study protocol and guidelines for COVID-19 standard care was based on a standard recommendation of the National and World Health Organization (WHO) (Methods S1).^7,14^

### PATIENT SELECTION AND PROCEDURES

A subset of 495 patients (age≥12 years) from the aforementioned patients were recruited for the reactogenicity and immunogenicity follow-up study after COVID-19 recovery at 21-150 days post COVID-19 onset to test for SARS-CoV-2 antibodies and a surrogate virus neutralization test (sVNT) against SARS-CoV-2 Wuhan and Delta variants (Method S1). This follow-up study was approved by the Institutional Review Board, Faculty of Medicine Siriraj Hospital, Mahidol University (COA: Si 833/2021, COA: Si 732/2021). At the study visit, all patients were classified into different exposure groups based on vaccination status prior to COVID-19 infection, study antibody, and PCR test (Method S1).

### OUTCOME MEASURES

Rationales for treatment of mild to moderate COVID-19 were shown in Method S1. In brief, the primary treatment strategy included early Favipiravir treatment. The primary composite outcome was to compare baseline clinical and biological characteristics of patients with Delta and Omicron variants of SARS-CoV-2 infections in the HI system. Treatment groups were categorized into 3 groups: (1) symptomatic treatment (S), (2) symptomatic treatment plus Favipiravir treatment (Favi), and (3) symptomatic treatment plus Favipiravir and dexamethasone treatment (Favi/Dexa). The date of disease onset was defined as the day when new-onset self-reported respiratory symptoms were observed. The durations of illness onset to first hospital admission, to first Favipiravir treatment, and to discharge up to 14 days were measured. Analyses considered viral load for comparisons of cycle threshold (Ct) values by vaccine exposure groups and self-reported symptoms. The Ct value of ≥30 was shown to correspond to 150 copies per milliliter or less, indicating low viral RNA.^15^

### SEROLOGIC ASSAYS

Patients were randomly invited to test for anti-SARS-CoV-2 receptor binding domain immunoglobulin G (anti-SARS-CoV-2 RBD IgG). A chemiluminescent microparticle immunoassay for qualitative detection of IgG against RBD of the SARS-CoV-2 spike protein and SARS-CoV-2 nucleocapsid protein (SARS-CoV-2 IgG II Quant for use with ARCHITECT; Abbott Laboratories, USA) were undertaken.^7^ Anti-SARS-CoV-2 RBD IgG assay was then converted to the WHO International Standard concentration as binding antibody unit (BAU) per milliliter.^16^ sVNT was undertaken against the original (Wuhan) strain and the Delta (B.1617.2) strain (Method S1).

### STATISTICAL ANALYSIS

Multivariable analysis was performed by binary logistic regression for vaccination variables. We used negative binomial mixed models to analyze factors associated with numeric variables including symptoms and the Chalder fatigue scale. Let *y*_*i*_ and *N*_*i*_ represented a number of symptoms or fatigue score and a number of persons time at risk *i*, (*i* = 1,…, *n*), respectively. We assumed that *y*_*i*_ conditional on *μ*_*i*_had the negative binomial distribution with mean *μ*_*i*_where *μ*_*i*_ = *N*_*i*_ exp(*ψ* _*i*_) was written as *y*_*i*_ | *μ*_*i*_ ∼ *NB*(*N*_*i*_ exp(*ψ* _*i*_)), where *ψ* _*i*_ = *X*_*i*_ ‘*β* + *v*_*i*_, *X* _*i*_’ represented the vector of space-specific covariates for a number of symptoms or a fatigue score and *β* as a vector of coefficients that were obtained by regression, *v*_*i*_ as a random effect with apprehending residual in area *i*, which was the unstructured heterogeneity that compiles effect of covariates in an unknown area. The area *i* had the relative risk as *r*_*i*_ = exp(*ψ* _*i*_). Then specification of the log-link function was log(*μ*_*i*_) = *X*_*i*_ ‘*β* + *v*_*i*_ + log(*N*_*i*_). Cox regression analysis was used to analyze the factors of the negative conversion time (NCT) of SARS-CoV-2 RNA. NCT is closely related to clinical manifestation and disease progression in COVID-19 patients. First, univariate analysis was performed and the indicators with statistical significance were analyzed for Kaplan-Meier survival analysis. A Cox proportional hazard model was used for multivariate analysis. Normally distributed continuous variables were summarized as the mean ± SD; otherwise, median (interquartile range, IQR) was used. Categorical variables were expressed using numbers and percentages. Statistical significance of Ct values, IgG, and sVNT were determined by Kruskal-Wallis and Dunn’s multiple comparisons test by using GraphPad Prism 9 (GraphPad Software, CA, USA). Other analyses were conducted using STATA version 17 (Stata Corp, TX, USA).

## RESULTS

### DEMOGRAPHIC AND CLINICAL INFORMATION

2704 and 2477 patients were enrolled between July-October 2021 and January-March 2022 during the Delta and Omicron pandemics, respectively (Table 1). Most patients were women (53.7-58.4%) and their mean age were younger in the Omicron compared with the Delta (31.3±12.3 vs. 33.8±11.6 years, p<0.001), suggesting that middle-aged people were more susceptible to infection with mild symptoms than other age groups. Comorbidities differed significantly between two pandemics, including prevalence of hypertension (p<0.001), obesity (p=0.009), and neurologic disease (p<0.001). Common initial symptoms were cough (60.7%) and low-grade fever (95.2%) in the Delta, whereas were asymptomatic (39.1%) and cough (47.7%) in the Omicron. The median duration from disease onset to HI admission was 5.1 (IQR=2.4) and 2.8 (IQR=1.6) days in the Delta and Omicron, respectively (p=0.021). The median peak viral RNA based on Ct values during the Omicron (19.0 [IQR=5.7]) was lower than the Delta (21.0 [IQR=7.8], p<0.001). No correlation was observed in Ct values and vaccination status, during Delta vs. Omicron pandemics (*ns*). Retrospective analysis revealed patients receiving dexamethasone treatment significantly had Ct levels below 20 during the Delta (p<0.001), which indicated that Ct levels were associated with disease severity in the Delta but not in the Omicron.

**Table 1.**
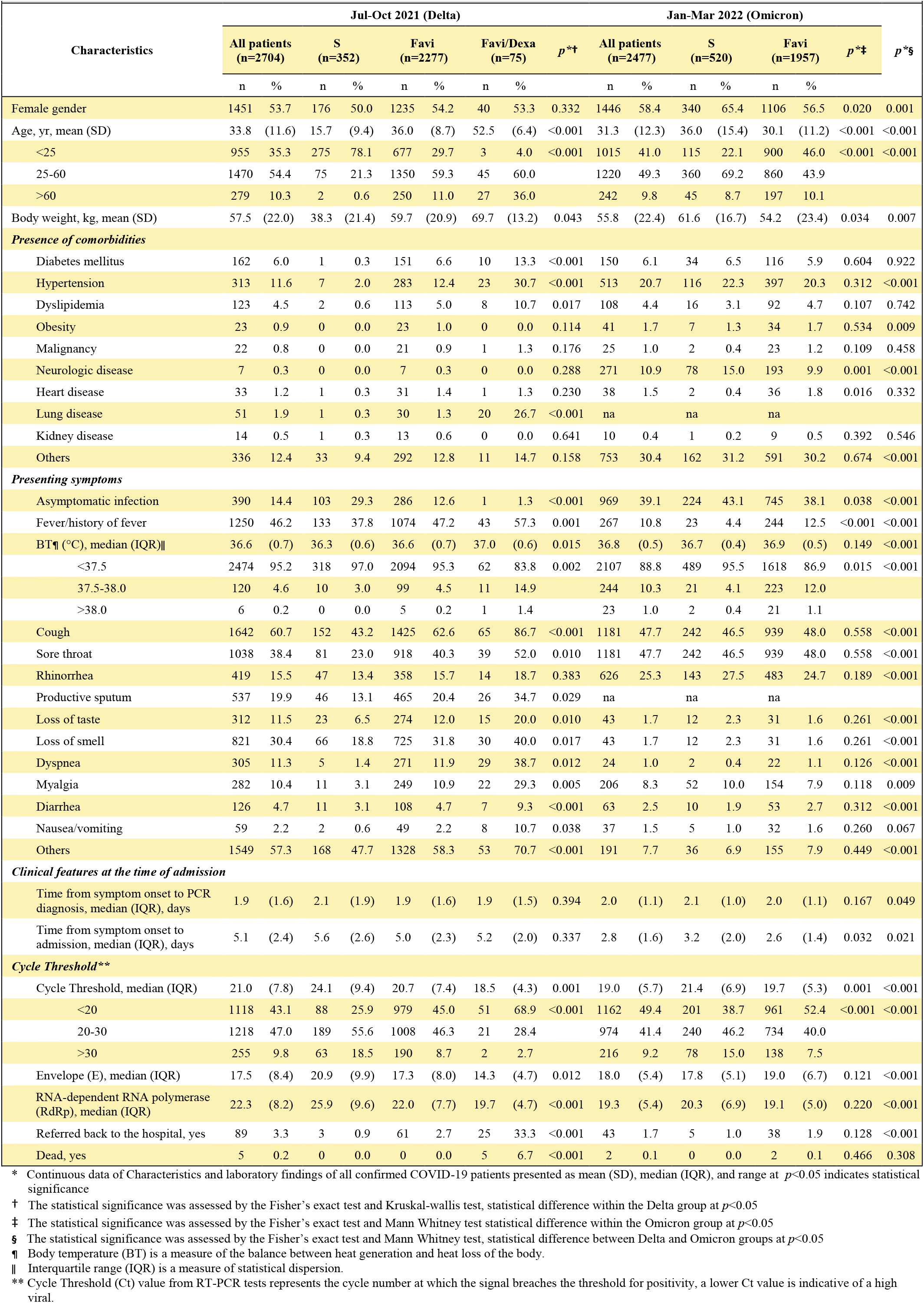
Characteristics and laboratory findings of all confirmed COVID-19 patients, and compared between those with symptomatic treatment (S), symptomatic treatment plus 5 - 14 days standard Favipiravir treatment (Favi) and symptomatic treatment plus 5 - 14 days standard Favipiravir treatment plus dexamethasone reatment (Favi/Dexa)*

### REHOSPITALIZED COVID-19 PATIENTS

Eighty-nine (3.3%) and forty-three (1.7%) patients in HI system were eventually referred back to the hospital during the Delta and Omicron, mostly due to worsening conditions, respectively, suggesting aggravated COVID-19 (Table 2). The mean age of patients in the Delta was older than the Omicron (55, IQR=24 vs. 33, IQR =14, p<0.001). More than a quarter of patients had underlying diseases, including diabetes (16.3-25.8%) and hypertension (23.3-31.5%). Compared with the Omicron, patients in the Delta had marked lymphocytopenia (0.4-fold) and neutrocytosis (1.8-fold), higher serum C-reactive protein (CRP) (21.2-fold), aspartate aminotransferase (1.6-fold), alanine aminotransferase (2.3-fold), and D-dimer (2-fold) (p<0.05). Vaccination with at least 2 doses was associated with reduced readmission rates in the Delta (OR=0.305, 95% CI=0.189-0.504) and the Omicron (OR=0.131, 95% CI, 0.052-0.334), as compared with unvaccinated and partial vaccination groups.

**Table 2.**
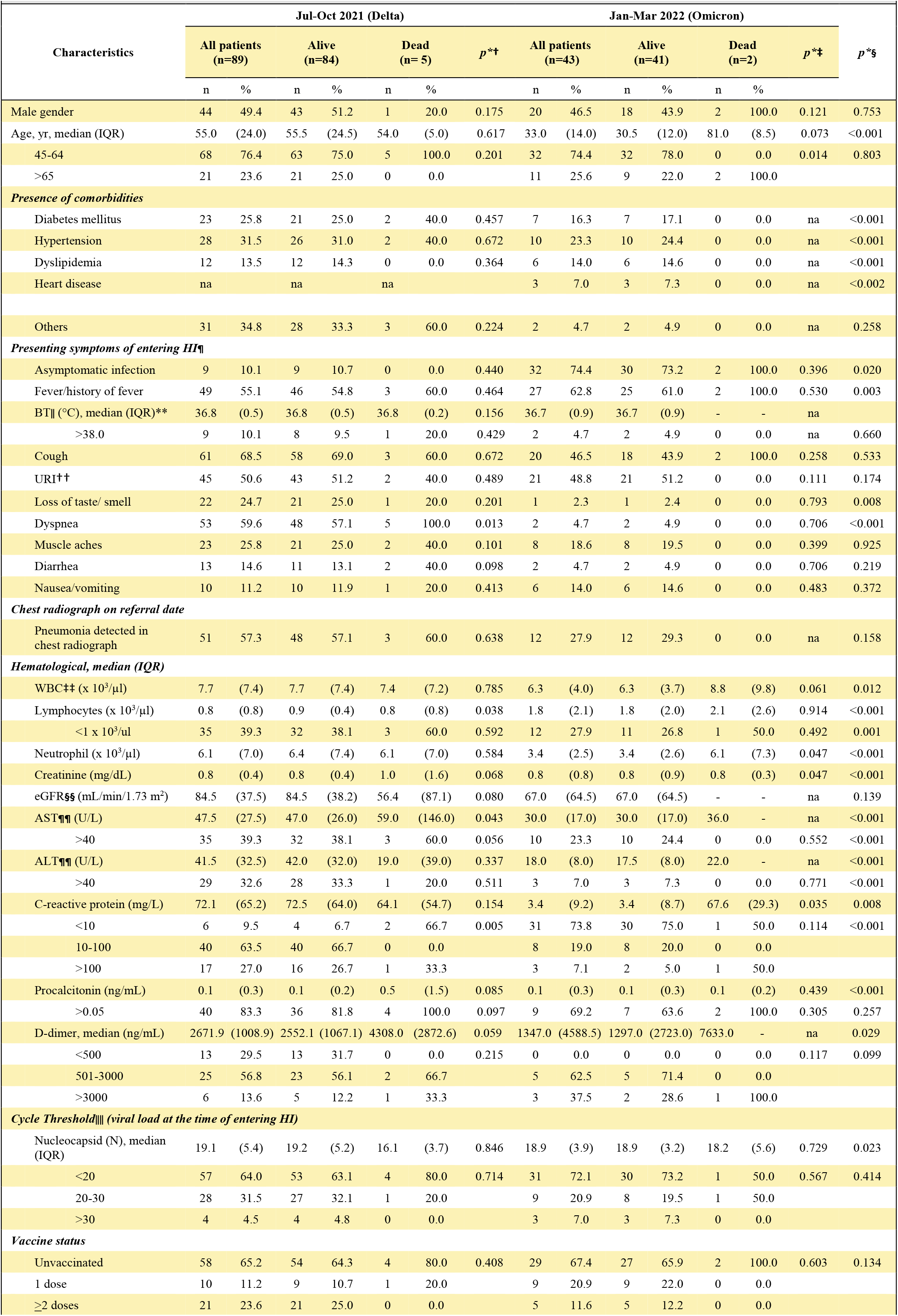
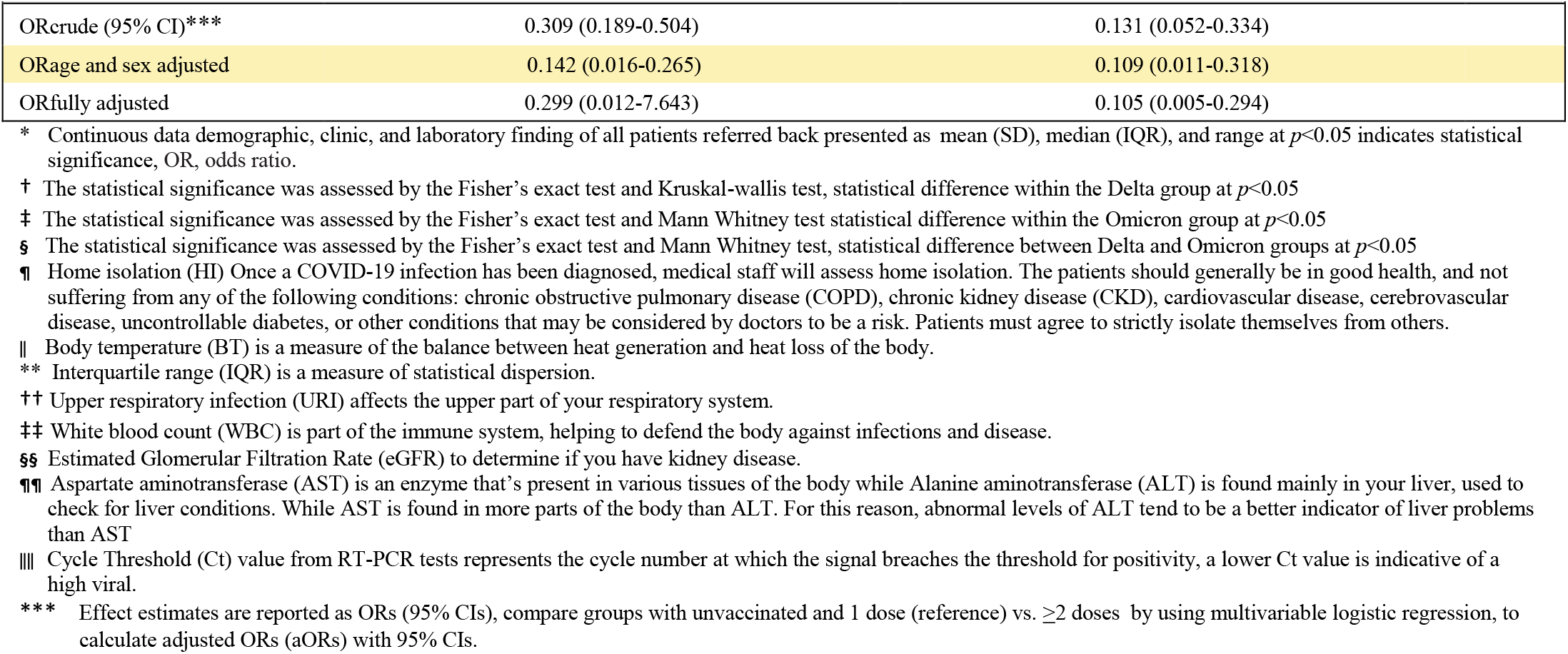
Demographic, clinic, and laboratory finding of all of patients referred back for in-patients care and compared between those during the Delta and the O micron pandemic*.

| Characteristics | Jul-Oct 2021 (Delta) |  |  |  |  |  |  | Jan-Mar 2022 (Omicron) |  |  |  |  |  |  | p*§ |
| --- | --- | --- | --- | --- | --- | --- | --- | --- | --- | --- | --- | --- | --- | --- | --- |
|  | All patients (n=89) |  | Alive (n=84) |  | Dead (n= 5) |  | p*† | All patients (n=43) |  | Alive (n=41) |  | Dead (n=2) |  | p*‡ |  |
|  | n | % | n | % | n | % |  | n | % | n | % | n | % |  |  |
| Male gender | 44 | 49.4 | 43 | 51.2 | 1 | 20.0 | 0.175 | 20 | 46.5 | 18 | 43.9 | 2 | 100.0 | 0.121 | 0.753 |
| Age, yr, median (IQR) | 55.0 | (24.0) | 55.5 | (24.5) | 54.0 | (5.0) | 0.617 | 33.0 | (14.0) | 30.5 | (12.0) | 81.0 | (8.5) | 0.073 | <0.001 |
| 45-64 | 68 | 76.4 | 63 | 75.0 | 5 | 100.0 | 0.201 | 32 | 74.4 | 32 | 78.0 | 0 | 0.0 | 0.014 | 0.803 |
| >65 | 21 | 23.6 | 21 | 25.0 | 0 | 0.0 |  | 11 | 25.6 | 9 | 22.0 | 2 | 100.0 |  |  |
| Presence of comorbidities |  |  |  |  |  |  |  |  |  |  |  |  |  |  |  |
| Diabetes mellitus | 23 | 25.8 | 21 | 25.0 | 2 | 40.0 | 0.457 | 7 | 16.3 | 7 | 17.1 | 0 | 0.0 | na | <0.001 |
| Hypertension | 28 | 31.5 | 26 | 31.0 | 2 | 40.0 | 0.672 | 10 | 23.3 | 10 | 24.4 | 0 | 0.0 | na | <0.001 |
| Dyslipidemia | 12 | 13.5 | 12 | 14.3 | 0 | 0.0 | 0.364 | 6 | 14.0 | 6 | 14.6 | 0 | 0.0 | na | <0.001 |
| Heart disease | na |  | na |  | na |  |  | 3 | 7.0 | 3 | 7.3 | 0 | 0.0 | na | <0.002 |
| Others | 31 | 34.8 | 28 | 33.3 | 3 | 60.0 | 0.224 | 2 | 4.7 | 2 | 4.9 | 0 | 0.0 | na | 0.258 |
| Presenting symptoms of entering HI¶ |  |  |  |  |  |  |  |  |  |  |  |  |  |  |  |
| Asymptomatic infection | 9 | 10.1 | 9 | 10.7 | 0 | 0.0 | 0.440 | 32 | 74.4 | 30 | 73.2 | 2 | 100.0 | 0.396 | 0.020 |
| Fever/history of fever | 49 | 55.1 | 46 | 54.8 | 3 | 60.0 | 0.464 | 27 | 62.8 | 25 | 61.0 | 2 | 100.0 | 0.530 | 0.003 |
| BT\|\| (°C), median (IQR)** | 36.8 | (0.5) | 36.8 | (0.5) | 36.8 | (0.2) | 0.156 | 36.7 | (0.9) | 36.7 | (0.9) | - | - | na |  |
| >38.0 | 9 | 10.1 | 8 | 9.5 | 1 | 20.0 | 0.429 | 2 | 4.7 | 2 | 4.9 | 0 | 0.0 | na | 0.660 |
| Cough | 61 | 68.5 | 58 | 69.0 | 3 | 60.0 | 0.672 | 20 | 46.5 | 18 | 43.9 | 2 | 100.0 | 0.258 | 0.533 |
| URI†† | 45 | 50.6 | 43 | 51.2 | 2 | 40.0 | 0.489 | 21 | 48.8 | 21 | 51.2 | 0 | 0.0 | 0.111 | 0.174 |
| Loss of taste/ smell | 22 | 24.7 | 21 | 25.0 | 1 | 20.0 | 0.201 | 1 | 2.3 | 1 | 2.4 | 0 | 0.0 | 0.793 | 0.008 |
| Dyspnea | 53 | 59.6 | 48 | 57.1 | 5 | 100.0 | 0.013 | 2 | 4.7 | 2 | 4.9 | 0 | 0.0 | 0.706 | <0.001 |
| Muscle aches | 23 | 25.8 | 21 | 25.0 | 2 | 40.0 | 0.101 | 8 | 18.6 | 8 | 19.5 | 0 | 0.0 | 0.399 | 0.925 |
| Diarrhea | 13 | 14.6 | 11 | 13.1 | 2 | 40.0 | 0.098 | 2 | 4.7 | 2 | 4.9 | 0 | 0.0 | 0.706 | 0.219 |
| Nausea/vomiting | 10 | 11.2 | 10 | 11.9 | 1 | 20.0 | 0.413 | 6 | 14.0 | 6 | 14.6 | 0 | 0.0 | 0.483 | 0.372 |
| Chest radiograph on referral date |  |  |  |  |  |  |  |  |  |  |  |  |  |  |  |
| Pneumonia detected in chest radiograph | 51 | 57.3 | 48 | 57.1 | 3 | 60.0 | 0.638 | 12 | 27.9 | 12 | 29.3 | 0 | 0.0 | na | 0.158 |
| Hematological, median (IQR) |  |  |  |  |  |  |  |  |  |  |  |  |  |  |  |
| WBC‡‡ (x 10<sup>3</sup>/μl) | 7.7 | (7.4) | 7.7 | (7.4) | 7.4 | (7.2) | 0.785 | 6.3 | (4.0) | 6.3 | (3.7) | 8.8 | (9.8) | 0.061 | 0.012 |
| Lymphocytes (x 10<sup>3</sup>/μl) | 0.8 | (0.8) | 0.9 | (0.4) | 0.8 | (0.8) | 0.038 | 1.8 | (2.1) | 1.8 | (2.0) | 2.1 | (2.6) | 0.914 | <0.001 |
| <1 x 10<sup>3</sup>/ul | 35 | 39.3 | 32 | 38.1 | 3 | 60.0 | 0.592 | 12 | 27.9 | 11 | 26.8 | 1 | 50.0 | 0.492 | 0.001 |
| Neutrophil (x 10<sup>3</sup>/μl) | 6.1 | (7.0) | 6.4 | (7.4) | 6.1 | (7.0) | 0.584 | 3.4 | (2.5) | 3.4 | (2.6) | 6.1 | (7.3) | 0.047 | <0.001 |
| Creatinine (mg/dL) | 0.8 | (0.4) | 0.8 | (0.4) | 1.0 | (1.6) | 0.068 | 0.8 | (0.8) | 0.8 | (0.9) | 0.8 | (0.3) | 0.047 | <0.001 |
| eGFR§§ (mL/min/1.73 m<sup>2</sup>) | 84.5 | (37.5) | 84.5 | (38.2) | 56.4 | (87.1) | 0.080 | 67.0 | (64.5) | 67.0 | (64.5) | - | - | na | 0.139 |
| AST¶¶ (U/L) | 47.5 | (27.5) | 47.0 | (26.0) | 59.0 | (146.0) | 0.043 | 30.0 | (17.0) | 30.0 | (17.0) | 36.0 | - | na | <0.001 |
| >40 | 35 | 39.3 | 32 | 38.1 | 3 | 60.0 | 0.056 | 10 | 23.3 | 10 | 24.4 | 0 | 0.0 | 0.552 | <0.001 |
| ALT¶¶ (U/L) | 41.5 | (32.5) | 42.0 | (32.0) | 19.0 | (39.0) | 0.337 | 18.0 | (8.0) | 17.5 | (8.0) | 22.0 | - | na | <0.001 |
| >40 | 29 | 32.6 | 28 | 33.3 | 1 | 20.0 | 0.511 | 3 | 7.0 | 3 | 7.3 | 0 | 0.0 | 0.771 | <0.001 |
| C-reactive protein (mg/L) | 72.1 | (65.2) | 72.5 | (64.0) | 64.1 | (54.7) | 0.154 | 3.4 | (9.2) | 3.4 | (8.7) | 67.6 | (29.3) | 0.035 | 0.008 |
| <10 | 6 | 9.5 | 4 | 6.7 | 2 | 66.7 | 0.005 | 31 | 73.8 | 30 | 75.0 | 1 | 50.0 | 0.114 | <0.001 |
| 10-100 | 40 | 63.5 | 40 | 66.7 | 0 | 0.0 |  | 8 | 19.0 | 8 | 20.0 | 0 | 0.0 |  |  |
| >100 | 17 | 27.0 | 16 | 26.7 | 1 | 33.3 |  | 3 | 7.1 | 2 | 5.0 | 1 | 50.0 |  |  |
| Procalcitonin (ng/mL) | 0.1 | (0.3) | 0.1 | (0.2) | 0.5 | (1.5) | 0.085 | 0.1 | (0.3) | 0.1 | (0.3) | 0.1 | (0.2) | 0.439 | <0.001 |
| >0.05 | 40 | 83.3 | 36 | 81.8 | 4 | 100.0 | 0.097 | 9 | 69.2 | 7 | 63.6 | 2 | 100.0 | 0.305 | 0.257 |
| D-dimer, median (ng/mL) | 2671.9 | (1008.9) | 2552.1 | (1067.1) | 4308.0 | (2872.6) | 0.059 | 1347.0 | (4588.5) | 1297.0 | (2723.0) | 7633.0 | - | na | 0.029 |
| <500 | 13 | 29.5 | 13 | 31.7 | 0 | 0.0 | 0.215 | 0 | 0.0 | 0 | 0.0 | 0 | 0.0 | 0.117 | 0.099 |
| 501-3000 | 25 | 56.8 | 23 | 56.1 | 2 | 66.7 |  | 5 | 62.5 | 5 | 71.4 | 0 | 0.0 |  |  |
| >3000 | 6 | 13.6 | 5 | 12.2 | 1 | 33.3 |  | 3 | 37.5 | 2 | 28.6 | 1 | 100.0 |  |  |
| Cycle Threshold‖‖ (viral load at the time of entering HI) |  |  |  |  |  |  |  |  |  |  |  |  |  |  |  |
| Nucleocapsid (N), median (IQR) | 19.1 | (5.4) | 19.2 | (5.2) | 16.1 | (3.7) | 0.846 | 18.9 | (3.9) | 18.9 | (3.2) | 18.2 | (5.6) | 0.729 | 0.023 |
| <20 | 57 | 64.0 | 53 | 63.1 | 4 | 80.0 | 0.714 | 31 | 72.1 | 30 | 73.2 | 1 | 50.0 | 0.567 | 0.414 |
| 20-30 | 28 | 31.5 | 27 | 32.1 | 1 | 20.0 |  | 9 | 20.9 | 8 | 19.5 | 1 | 50.0 |  |  |
| >30 | 4 | 4.5 | 4 | 4.8 | 0 | 0.0 |  | 3 | 7.0 | 3 | 7.3 | 0 | 0.0 |  |  |
| Vaccine status |  |  |  |  |  |  |  |  |  |  |  |  |  |  |  |
| Unvaccinated | 58 | 65.2 | 54 | 64.3 | 4 | 80.0 | 0.408 | 29 | 67.4 | 27 | 65.9 | 2 | 100.0 | 0.603 | 0.134 |
| 1 dose | 10 | 11.2 | 9 | 10.7 | 1 | 20.0 |  | 9 | 20.9 | 9 | 22.0 | 0 | 0.0 |  |  |
| ≥2 doses | 21 | 23.6 | 21 | 25.0 | 0 | 0.0 |  | 5 | 11.6 | 5 | 12.2 | 0 | 0.0 |  |  |

|  |  |  |
| --- | --- | --- |
| ORcrude (95% CI)*** | 0.309 (0.189-0.504) | 0.131 (0.052-0.334) |
| ORage and sex adjusted | 0.142 (0.016-0.265) | 0.109 (0.011-0.318) |
| ORfully adjusted | 0.299 (0.012-7.643) | 0.105 (0.005-0.294) |

\* Continuous data demographic, clinic, and laboratory finding of all patients referred back presented as mean (SD), median (IQR), and range at $p < 0.05$ indicates statistical significance, OR, odds ratio.
† The statistical significance was assessed by the Fisher's exact test and Kruskal-wallis test, statistical difference within the Delta group at $p < 0.05$
‡ The statistical significance was assessed by the Fisher's exact test and Mann Whitney test statistical difference within the Omicron group at $p < 0.05$
§ The statistical significance was assessed by the Fisher's exact test and Mann Whitney test, statistical difference between Delta and Omicron groups at $p < 0.05$
¶ Home isolation (HI) Once a COVID-19 infection has been diagnosed, medical staff will assess home isolation. The patients should generally be in good health, and not suffering from any of the following conditions: chronic obstructive pulmonary disease (COPD), chronic kidney disease (CKD), cardiovascular disease, cerebrovascular disease, uncontrollable diabetes, or other conditions that may be considered by doctors to be a risk. Patients must agree to strictly isolate themselves from others.
|| Body temperature (BT) is a measure of the balance between heat generation and heat loss of the body.
\*\* Interquartile range (IQR) is a measure of statistical dispersion.
†† Upper respiratory infection (URI) affects the upper part of your respiratory system.
‡‡ White blood count (WBC) is part of the immune system, helping to defend the body against infections and disease.
§§ Estimated Glomerular Filtration Rate (eGFR) to determine if you have kidney disease.
¶¶ Aspartate aminotransferase (AST) is an enzyme that's present in various tissues of the body while Alanine aminotransferase (ALT) is found mainly in your liver, used to check for liver conditions. While AST is found in more parts of the body than ALT. For this reason, abnormal levels of ALT tend to be a better indicator of liver problems than AST
|||| Cycle Threshold (Ct) value from RT-PCR tests represents the cycle number at which the signal breaches the threshold for positivity, a lower Ct value is indicative of a high viral.
\*\*\* Effect estimates are reported as ORs (95% CIs), compare groups with unvaccinated and 1 dose (reference) vs. $\geq 2$ doses by using multivariable logistic regression, to calculate adjusted ORs (aORs) with 95% CIs.

### NUMBERS OF SYMPTOMS AND A FATIGUE SCORE DURING HI ADMISSION

The risk factors, including gender, severity of illness, and vaccination status, were significantly related to an increased fatigue symptom as determined by the Chalder fatigue scale^16^ (Table 3 and Table S1). Neutralizing antibody titers were independently associated with the number of symptoms (RR= 1.22, 95% CI, 1.05-1.42, p=0.009). However, the association of neutralizing antibody titers was not statistically significant for the fatigue. The severity of initial illness was associated with the persistent fatigue (aRR=1.43, 95% CI, 1.37-1.72, p=0.039) and weakly associated with the number of symptoms (aRR=1.22, 95% CI, 1.02-1.45, p=0.03). In the stratified analysis of the patients, increased antibody titers remained associated with the number of symptoms (aRR=1.04, 95% CI, 1.01-1.08, p=0.025) and the fatigue scale (aRR=1.09, 95% CI, 1.02-1.16, p=0.015). Patients who were vaccinated prior to COVID-19 infection reported a significantly lower number of symptoms (p<0.001) and the fatigue score (p=0.01) than unvaccinated patients. To analyze factors on NCT of SARS-CoV-2 RNA systematically, the multivariate Cox’s proportional hazard model was used to analyze the multivariate analyses. We revealed that fever (Exp(B), 0.75; 95% CI, 0.70-0.80, p<0.001), cough (Exp(B), 0.84; 95% CI, 0.81-0.87, p<0.001) and loss of smell (Exp(B), 0.81; 95% CI, 0.77-0.85, p<0.001) were independent risk factors of prolonged NCT of SARS-CoV-2 RNA in patients with COVID-19 (Table S6).

**Table 3.** COVID-19 patient factors associated with increasing number of symptoms and higher fatigue score at 14-days follow-up—negative binomial mixed models (July 2 021 to March 2022)*

| Characteristics | n % |  | Number of symptoms<sup>†</sup> (0-12) |  |  |  |  |  | Fatigue score<sup>‡</sup> (0-27) |  |  |  |  |  |
| --- | --- | --- | --- | --- | --- | --- | --- | --- | --- | --- | --- | --- | --- | --- |
|  |  |  | RR<sup>§</sup> | 95%CI<sup>¶</sup> | *p*<sup>**</sup> | aRR<sup>**</sup> | 95%CI | *p*<sup>**</sup> | RR | 95%CI | *p*<sup>††</sup> | aRR | 95%CI | *p*<sup>††</sup> |
| Female gender | 2194 | 58.4 | 1.02 | (0.94, 1.10) | 0.650 | 1.03 | (0.96, 1.11) | 0.361 | 1.13 | (0.99, 1.30) | 0.071 | 1.15 | (1.00, 1.31) | 0.049* |
| Age, yr, median (range) | 31 | (17-47) | 1.00 | (1.00, 1.00) | 0.966 | 1.00 | (1.00, 1.00) | 0.146 | 1.00 | (0.99, 1.00) | 0.458 | 1.00 | (0.99, 1.00) | 0.292 |
| **Comorbidity** |  |  |  |  |  |  |  |  |  |  |  |  |  |  |
| Hypertension | 769 | 20.5 | 1.05 | (0.95, 1.16) | 0.322 | 1.04 | (0.93, 1.16) | 0.488 | 0.97 | (0.81, 1.15) | 0.708 | - | -- | - |
| Dyslipidemia | 231 | 6.2 | 1.01 | (0.89, 1.15) | 0.890 | 1.07 | (0.93, 1.22) | 0.353 | 0.97 | (0.77, 1.22) | 0.781 | 1.11 | (0.86, 1.42) | 0.427 |
| Diabetes mellitus | 311 | 8.3 | 0.99 | (0.87, 1.13) | 0.914 | 0.95 | (0.83, 1.09) | 0.491 | 0.83 | (0.65, 1.05) | 0.115 | 0.81 | (0.63, 1.04) | 0.104 |
| Asthma/COPD<sup>‡‡</sup> | 97 | 4.8 | 0.99 | (0.85, 1.15) | 0.882 | 0.99 | (0.86, 1.14) | 0.882 | 0.84 | (0.64, 1.10) | 0.211 | 0.85 | (0.65, 1.11) | 0.232 |
| Chronic heart disease | 65 | 1.7 | 1.07 | (0.83, 1.39) | 0.592 | 1.07 | (0.84, 1.37) | 0.568 | 1.22 | (0.77, 1.93) | 0.39 | 1.30 | (0.83, 2.04) | 0.252 |
| Severity of initial illness | 102 | 2.7 | 1.34 | (1.11, 1.61) | 0.002* | 1.22 | (1.02, 1.45) | 0.030* | 1.46 | (0.95, 1.75) | 0.051 | 1.43 | (1.37, 1.72) | 0.039* |
| Immunosuppression<sup>§§</sup>, median (range) | 8.89 | (7.94-9.53) | 1.05 | (1.03, 1.08) | <0.001* | 1.04 | (1.01, 1.08) | 0.025* | 1.04 | (1.00, 1.10) | 0.074 | 1.09 | (1.02, 1.16) | 0.015* |
| Neutralizing antibody titers<sup>¶¶</sup>, median (range) | 4.58 | (4.55-4.59) | 1.22 | (1.08, 1.37) | 0.001* | 1.22 | (1.05, 1.42) | 0.009* | 1.18 | (0.80, 1.20) | 0.876 | 1.14 | (0.66, 1.19) | 0.324 |
| Vaccinated, yes | 2577 | 68.6 | 0.81 | (0.76, 0.87) | <0.001* | 0.78 | (0.72, 0.84) | <0.001* | 0.85 | (0.75, 0.97) | 0.014* | 0.84 | (0.73, 0.96) | 0.011* |

\* Analysis of associated factors was done by negative binomial mixed models. aRR, adjusted rate ratio; RR, rate ratio. Statistical significance at the level of $p < 0.05$ is shown in bold text.
<sup>†</sup> Patients were assessed for 12 symptoms mentioned in Table S1.
<sup>‡</sup> Chalder fatigue score is validated only for patients aged >18 years (n=3756); possible fatigue scores range from 0 (no fatigue) to 27 (worst possible fatigue).
<sup>§</sup> The relative risk (RR) is the risk of the event in an experimental group relative to that in a control group.
<sup>¶</sup> 95% confidence interval (CI) is used to estimate the precision of the OR
<sup>||</sup> Factors with significance level $p < 0.1$ in univariable analysis were included in the multivariable analysis of symptoms at 14-days follow-up.
<sup>\*\*</sup> Adjusted rate ratio (aRR) is the difference in increased risk of symptoms and fatigue score.
<sup>††</sup> Factors with significance level $p < 0.1$ in univariable analysis were included in the multivariable analysis of fatigue score at 14-days follow-up.
<sup>‡‡</sup> COPD is chronic obstructive pulmonary disease.
<sup>§§</sup> SARS-CoV-2 spike protein antibody titers, log<sub>10</sub> transformed.
<sup>¶¶</sup> Neutralizing antibody titers, log<sub>10</sub> transformed.

### CLINICAL MANIFESTATION AND VIRAL BURDEN

Higher peak Ct values were slightly prevalent with numbers of doses of ChAdOx1 or CoronaVac when compared the Delta with the Omicron-dominant, and strongly decreased in those unvaccinated individuals in the Omicron (19 [IQR=17-22]) as compared with the Delta-dominant (21 [IQR=18-26]) age/gender-adjusted trend-*p*<0.001, Figure 1A, 1B). No difference was observed from those vaccinated with either ChAdOx1 or CoronaVac (age/gender-adjusted *p*=0.175) indicating that vaccination was still valuable in reducing viral load. Considering effect of pandemic waves (Delta vs. Omicron), new PCR-positives were likely to be in the low Ct subpopulation in most vaccinations, regardless of doses, types of vaccines and time since the last vaccination, during the Omicron while Ct levels tended to be varied during the Delta (Figure 1C-1I). In the Delta pandemic, BioNTech and Moderna vaccines were not available in Thailand, so we did not have Ct values related to these vaccines (Figure 1C-1D). During the Delta but not the Omicron pandemics, patients who had at least two-dose vaccination prior to COVID-19 infection reported a significantly lower number and probability of any symptoms (OR=0.25, 95% CI, 0.12-0.52, *p*<0.001) and common COVID-19 symptoms (cough, fever, anosmia/ageusia, [OR=0.28, 95% CI=0.13 - 0.58, *p*<0.001]) than unvaccinated individuals (Figure 1J, Table S4). No correlation between Ct values and probability of reporting any symptoms was noted in the Omicron pandemic (Figure 1K-1N, Table S5).

**Figure 1:**
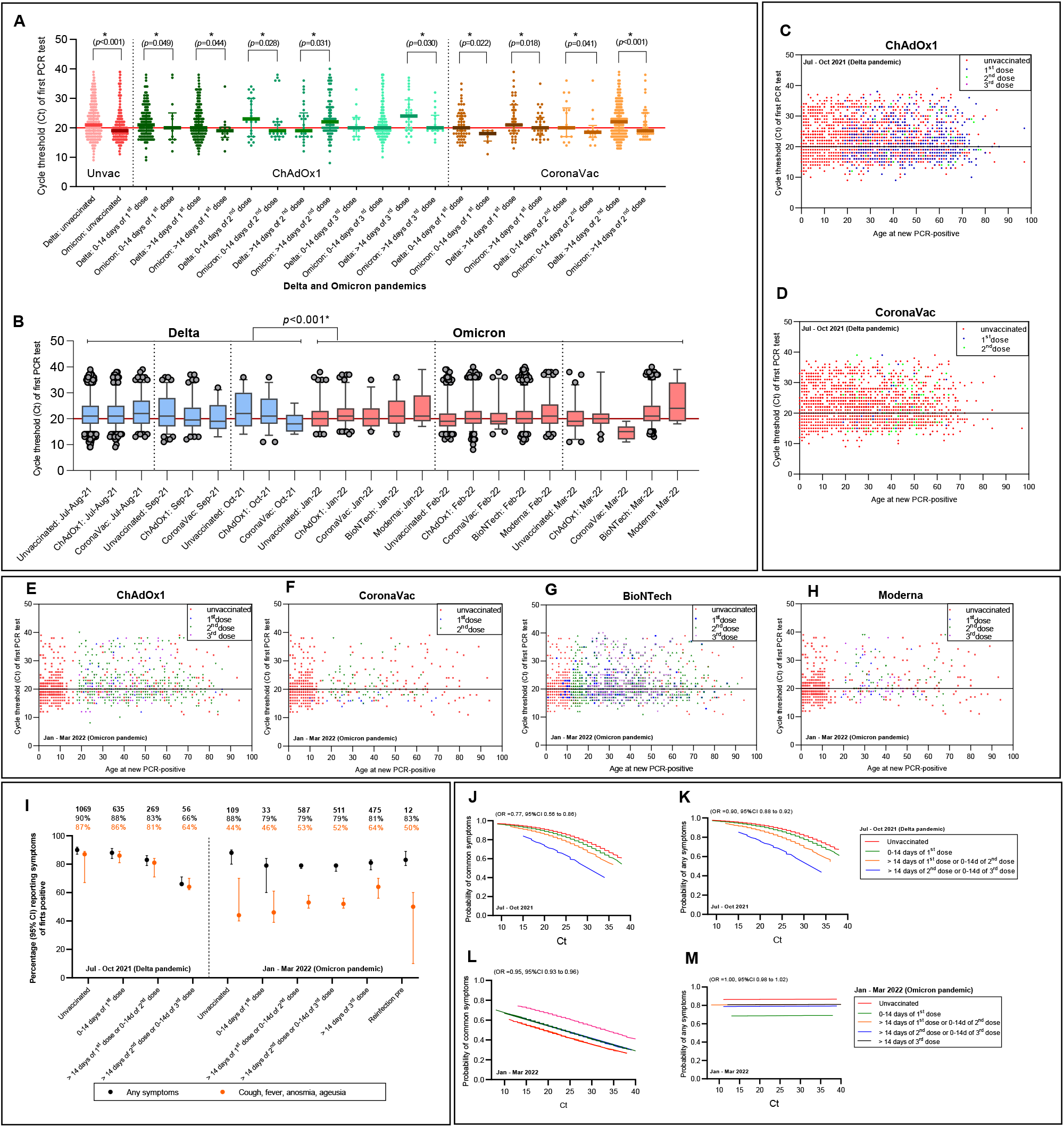
Ct value trajectories of confirmed COVID-19 infection and symptoms during the Delta and Omicron pandemics in vaccinated and unvaccinated individuals. (A) Ct values by vaccination/reinfection status and (B) Ct values by waves and vaccination type. (C-H) Ct values in PCR-positives after receiving ChAdOx1 or CoronaVac or BioNTech or Moderna vaccines or unvaccinated, regardless of vaccine doses by time since July 2021. Red dots are represented in all figures as reference values. (I) Self-report symptoms in PCR-positives by numbers of vaccination/reinfection status. Probability of reporting common (J, L) fever, cough, anosmia, ageusia or (K, M) any symptoms by Ct values and vaccination status in PCR-positives. \**p* <0.05

### IMMUNE RESPONSES AGAINST SARS-COV-2 VARIANTS

Higher antibody titers were observed in both unvaccinated COVID-19 and breakthrough COVID-19 patients vaccinated with both CoronaVac-prime or ChAdOx1-prime, regardless of numbers of doses, which reached their peak around 2-3 months post-COVID-19 (PC) and decreased gradually after 3 months. The RBD-IgG geometric mean titers (GMT) at baseline (1-2 months PC) were higher in the ChAdOx1 groups (1,2,3 doses) than in the CoronaVac groups but no different titers were observed between the two groups after 3 months PC (ChAdOx1: 1 dose [822 BAU/mL, 95% CI, 626-1081], 2 doses [945 BAU/mL, 95% CI, 567-1575], and 3 doses [886 BAU/mL, 95% CI, 588-1335] vs. (CoronaVac: 1 dose [1174 BAU/mL, 95% CI, 638-2158], and 2 doses [974 BAU/mL, 95% CI, 684-13786]). However, participants with a breakthrough infection had higher antibody titers at all time points compared with previously unvaccinated participants with COVID-19 infection (p<0.05, Figure 2A, Table S8). The GMT of anti-RBD IgG was significantly higher in unvaccinated after two months in both males and females (Figure 2G). Older individuals had significantly higher GMT of Anti-RBD IgG than the younger individuals in both unvaccinated and ChAdOx1 groups. Although there was no significant difference in anti-RBD IgG between age groups in the CoronaVac groups, anti-RBD IgG tended to be higher in the older individuals than the younger individuals (Figure 2L).

**Figure 2:**
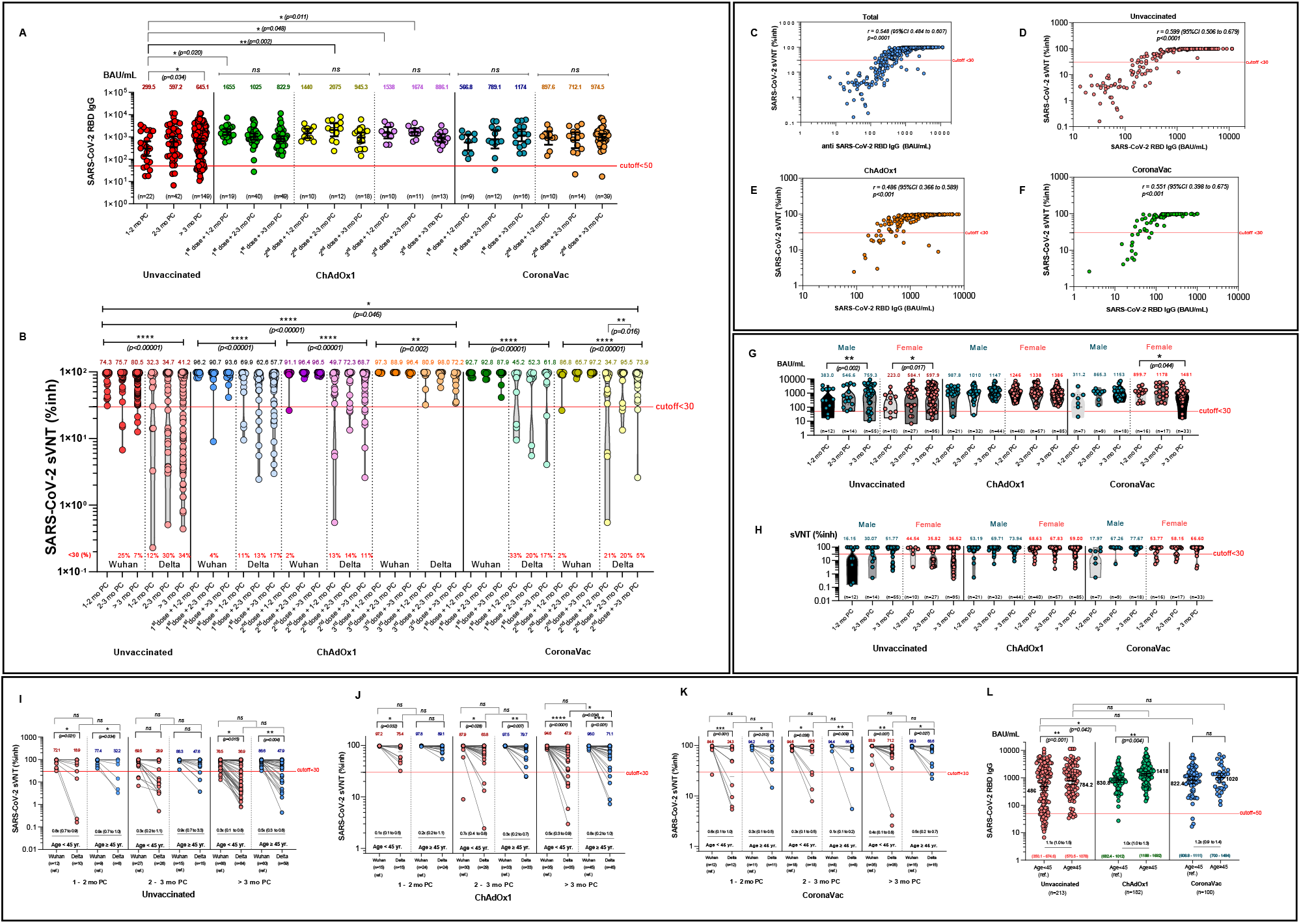
Immune responses after breakthrough COVID-19 infection with prior CoronaVac or ChAdOx1 vaccination during the Delta pandemic. (A) The scatter plot of geometric mean titers (GMT) of SARS-CoV-2 anti-spike protein receptor-binding domain antibodies (Anti-RBD IgG) concentrations in serum samples obtained from subjects after COVID-19 infection and with prior various vaccination status (CoronaVac vs. ChAdOx1). Sera at different time points from patients recovered from the COVID-19 are shown as reference level (red). (B) Scatter plots demonstrates an inhibition rate of Wuhan and Delta RBD-blocking antibodies measured using a surrogate viral neutralization test (sVNT) by vaccination/reinfection status, the lower dot line represents the cut-off level for seropositivity. All sera were from the patients during the Delta pandemic. (C-F) Dot plots show the correlation between the level of anti-SARS-CoV-2 RBD IgG and surrogate viral neutralization test (sVNT) for the SARS-CoV-2 delta variant in plasma of study participants (total, C) who were unvaccinated (D), or completed two doses of ChAdOxX1 (E), CoronaVac (F) and having breakthrough infection. (G, H) Anti-RBD IgG concentrations, sVNT by gender/vaccination/reinfection status, and sVNT (I-K) by age groups/vaccination/infection status and (L) antibodies (Anti-RBD IgG) by age/vaccination. \**p*<0.05; \*\**p*<0.01; \*\*\**p*<0.001; \*\*\*\**p*<0.0001.

Most patients had highly positive sVNT against Wuhan and Delta. Higher sVNT was observed mostly in breakthrough COVID-19 patients vaccinated with either CoronaVac-prime or ChAdOx1-prime, regardless of numbers of doses, which reached their peak around 2-3 months PC and decreased approximately 10-20% after 3 months as compared with 2-3 months PC (Figure 2B). Additionally, the titers were significantly higher against Wuhan as compared with the Delta variant (p<0.001). Using sVNT, the proportion of individuals with neutralizing test against Delta above sVNT cutoff of 30 was ≈66-88% of unvaccinated participants (pink dots), as compared with ≈83-89% of 1-2 doses of ChAdOx1 (pale blue dots), 100% of 3 doses of ChAdOx1, 67-95% of 1-2 doses of CoronaVac.

In fully-vaccinated individuals at 2-3 months PC, mean sVNT to the Delta relative to the Wuhan virus was reduced 0.7-fold (96.4→72.3, ChAdOx1 group). As compared with unvaccinated COVID-19, sVNT for the Delta variant of ChAdOx1-boosted individuals at 2-3 months PC were increased 2.8-fold (34.7→98) and 2.1-fold (34.7→72.3 when compared with 2-dose ChAdOx1 group) (Figure 2B). The anti-SARS-CoV-2 RBD IgG levels and sVNT against Delta variant were markedly correlated (r=0.486 to r=0.599) particularly in the unvaccinated group and vaccinated group (Figure 2C-2F). The proportion of plasma samples exhibiting such a neutralizing activity against Delta tended to be nearly 1-2-fold higher among the older than the younger individuals (Figure 2I-2K).

## DISCUSSION

At the time of our study, few studies had been published regarding the outcomes of mild-to-moderate COVID-19 in an outpatient setting. We affirmed the following important findings. First, in the majority of patients, illness is mild, and medical intervention is not needed, particularly in fully vaccinated individuals. This confirms that early access to treatment along with prompt responses from telehealth visits and anti-viral medication provides statistically favorable efficacy in sustaining COVID-19 and improving outcomes in an appropriate outpatient setting.^19^ Recent studies in a “hospitel” (a field hospital) demonstrated that patients receiving Favipiravir exhibited higher viral clearance rates than patients receiving standard symptomatic treatment and prevented hospitalization.^19^ A systematic review and meta-analysis of clinical trials summarized that Favipiravir exerted low efficacy in the term of reduced mortality in patients with mild to moderate COVID-19. But they pointed out that that might be due to treatment in many trials was delayed.^20^

Second, understanding the relationship between symptoms, viral load, disease severity and predictive immunity is crucial to planning for the next steps in the booster vaccination program. Our results are in contrast with a recent study^17^ reporting lower Ct values, for patients infected during mild Omicron pandemic, compared to patients infected during Delta pandemic. In the mild-to-moderate COVID-19 during the Delta and Omicron pandemics, antibodies including IgG and surrogate neutralizing titers are higher in patients with more severe common COVID-19 symptoms, associated with high viral load (lower Ct levels) and in older individuals who are generally vaccinated and have more severe symptoms than asymptomatic individuals.^5^ IgG and sVNT remained stable for at least 3 months. However, receiving booster vaccines will ensure better predictive immunity against COVID-19. In the Omicron pandemic, viral load was not correlated with symptoms. This was likely due to Omicron’s milder conditions, an improved vaccination campaign and quick access to medication treatment. Therefore, other variables should be considered for the assessment of symptoms. Our results are consistent with a previous study by Servellita *et al*.^5^ that examined neutralizing responses in Delta and Omicron breakthrough infections and displayed strong increases in antibody titers to Wuhan and Delta especially from boosting and added that in symptomatic or mild Delta and Omicron breakthrough infections, the extent of conferred cross-neutralizing immunity against Omicron and Delta is limited. However, Wratil *et al*.^18^ found that sera from patients with Omicron breakthrough infections enhanced Omicron viral neutralization significantly (17.4-fold).

It is well documented that COVID-19 is not only primarily manifested as a respiratory tract infection but also, emerging data indicate that it involves multiple systems. Several hematological laboratory investigations into lymphocytes, neutrophils and hemostasis including CRP, elevated D-dimer were altered significantly in COVID-19 patients, suggesting this can be regards as a potential indicator for both disease progression and effectiveness of therapy.^19^ Evidence showed that mild COVID-19 can be associated with a potent initial innate antiviral response induction and viral neutralization and might evade the host innate immune activation and consequently increase proinflammatory response and immune cell infiltration.^20,21^ Even though we did not have a complete set of these parameters for every subject, we fortunately had a subgroup of patients with worsening conditions who were eventually hospitalized and had their blood examined. Our results indicated that during the Delta pandemic these patients had increased neutrophils and lymphocytopenia, activation of the coagulation cascade. Although we did not observe this trend in patients during the Omicron pandemic, there was report that the Omicron COVID-19 patients had abnormal levels of neutrophils, lymphocytes, and monocytes and signs of coagulopath.^21-23^ Still, more in-depth research on the underlying etiology is necessary. A recent genome-wide association analysis (GWAS)^24^ showed associations of loci on chromosomes 5q32 and 9q21.13 with COVID-19 susceptibility and two suggestive loci on chromosomes 12q22 and 3p24.3 severity. Interestingly, the association signal on chromosome 5q32 coincided with *IL17B* encoding T cell-derived cytokine known as interleukin-17B (IL-17B). IL-17B was reported to play a role as a proinflammatory inducer in inflammatory disease, stimulating the release of tumor necrosis factor-α (TNF-α) and interleukin-1β (IL-1β) from a monocytic cell line resulting in neutrophil infiltration.^25,26^ This supports our finding of hyperneutrophilia seen in our COVID-19 cohort.

These data combined with ours could suggest that higher infectivity of Omicron may be related to a decreased viral load, probably lower past protective immunity against Omicron either from vaccines, or natural infection (Delta) and an asymptomatic stage of infected individuals with respiratory symptoms, age.^11^ But we found no meaningful difference in types of vaccines, numbers of vaccines or duration of disease symptoms of the Omicron variant as compared with the Delta variant. Still, substantial variation in patients’ symptoms and immunogenicity underscores the heterogenicity of protective immunity against future infections. However, an individual previously infected with SARS-Cov-2 is advised to receive a full vaccination or at least one additional dose of a vaccine after the infection to protect against reinfection from some circulating variants.^27^ High vaccination rates also help to reduce the transmission of COVID-19. However, vaccination rates are still low in some provinces, important risk groups, and low-income countries.^28^

There are several limitations to our study. We only have blood test from a small subset of hospitalized patients and assumed that these findings might be similar to all milder infected cases without hospitalization. At the time of analysis, we didn’t have serology data from patients during the Omicron pandemic and long term follow up data, so we could not determine the antibody levels against the Omicron variants and vaccine efficacy after the COVID-19 infection and its impact on the long COVID-19. Our future COVID-19 direction is to further gain insight into the long-term monitoring of neutralizing antibodies and to study if the breakthrough Omicron infection will have a protective immunity against reinfection of SARS-CoV-2 Omicron sub-lineages BA.4 and BA.5.

## CONCLUSION

Our study displayed important knowledge about immunogenicity and dynamics of the immune responses induced by post-COVID-19 together with prior vaccination, which help improve protection against current and new emerging SARS-CoV-2 variants. Our data suggest that in countries where vaccines are limited, full vaccination of most prime or mixed vaccines, and a booster shot for individuals with risks might be enough to induce high levels of short-term immunity and prevent hospitalization and death, regardless of higher viral burden or symptoms, especially during the Omicron pandemic in the absence of novel variants.

## Data Availability

All data produced in the present study are available upon reasonable request to the authors.

## ACKNOWLEDGEMENTS

We thank the patients and their families at Siriraj Hospital without whom collecting and providing this aggregate data would not have been successful. We are grateful for the support of the COVID-19 teams: Population Health and Nutrition Research Group, Tassanee Narkdontri, Nipaporn Teerawattanapong, Worapanit Lerdsaeng from Siriraj Center of Research Excellence for Diabetes and Obesity Research (SiCORE-DO), Research department and Department of Immunology at Faculty of Medicine Siriraj Hospital, Mahidol University and Utane Runpanich from Department of Immunology, Faculty of Medicine Siriraj Hospital, Mahidol University for performing IgG assays, and Center of Research Excellence on Therapeutic Proteins and Antibody Engineering, Department of Parasitology, Faculty of Medicine Siriraj Hospital, Mahidol University for performing the surrogate virus neutralization test.

## CONTRIBUTIONS

**Members of Siriraj Population Health and Nutrition Research Group (SPHERE):** Sureeporn Pumeiam, Bonggochpass Pinsawas, Pichanun Mongkolsucharitkul, Apinya Surawit, Tanyaporn Pongkunakorn, Sophida Suta, Thamonwan Manosan, Suphawan Ophakas and Korapat Mayurasakorn

## AUTHORS CONTRIBUTION

P.M., A.S. and KM conceived and designed the manuscript, analyzed the data and drafted the manuscript. P.P., N.S., W.S., C.W., C.A., T.S., R.T., M.C., G.S., P.P., S.S., S.M., and SPHERE performed patient care and revised and reviewed the manuscript. P.M. and A.S. K.M. performed critical revision of the manuscript. W.T. performed IgG testing and provided data establishing testing. N.S. and A.T. performed sVNT testing. A.S. performed biostatistical analysis and reviewed. The corresponding author (K.M.) attests that all listed authors meet authorship criteria and that noothers meeting the criteria have been omitted. All staff for COVID-19 care were listed in the supplement appendix. All authors read the manuscript and agreed to its contents.

## DATA AVAILABILITY

Raw data used in this study, including de-identified patient metadata and test results, are available upon request.

