## Supplementary Appendix for "Immunity response against mild-to-moderate breakthrough COVID-19"

This appendix has been provided by the authors to give readers additional information about the work.

| **Contents:** | **Page** |
| --- | --- |
| Investigators and support team members | 3 |
| Figure S1: Study source recruitment and enrollment | 5 |
| Methods S1: Study design, eligibility, inclusion & exclusion criteria | 6 |
| Figure S2: Trend of SARS-CoV-2 variants circulating in Thailand, deaths and vaccination status during analysis period | 10 |
| Table S1: General comorbidity questionnaire | 11 |
| Table S2: Characteristics of participants at enrollment (Jul-Oct 2021 (Delta)) by age | 14 |
| Table S3: Characteristics of participants at enrollment (Jan-Mar 2022 (Omicron)) by age | 17 |
| Table S4: Characteristics of the first positive symptoms between the Delta and Omicron pandemics for people aged >18 years | 20 |
| Table S5: Independent associations with symptoms reporting between the Delta and Omicron pandemics for people aged >18 years | 21 |
| Table S6: Independent factors of NCT of SARS-CoV-2 RNA by the multivariate Cox’s proportional hazard model by age >18 years (Jul-Oct 2021 (Delta)) | 22 |
| Figure S3: Kaplan-Meier estimate of time to symptom alleviation as reported by Covid-19-adapted FLU-PRO questionnaire (Jul-Oct 2021 (Delta)) | 23 |
| Table S7: Characteristics of the participants and geometric mean humoral immunogenicity assay responses according to anti-SARS-CoV-2 nucleocapsid protein and vaccine group | 24 |
| Table S8: Anti-RBD IgG and sVNT geometric mean concentration (GMC) among unvaccinated, ChAdOx1, and CoronaVac | 26 |
| Table S9: Anti-RBD IgG and sVNT geometric mean concentration (GMC) among unvaccinated, ChAdOx1, and CoronaVac by age | 27 |
| Table S10: SARS-CoV-2 sVNT geometric mean concentration (GMC) among unvaccinated, ChAdOx1, and CoronaVac by Wuhan and Delta variants | 28 |
| Figure S4: Plasma Anti-Immunoglobulin G (IgG) and Antibody (sVNT) levels against SARS-Cov-2 | 29 |

**Investigators and Support Team Members**

**SI-Home Study Team**

This project authors gratefully express our enormous gratitude to all members of SPHERE staff members and research assistants as well as all staff of Siriraj Hospital who work tirelessly to take care of individuals with COVID-19. This study and work done would not have been finished without their dedications and efforts. We also would like to acknowledge Associate Professor. Visit Vamvanij, the director of Siriraj Hospital for his guidance for initiation of this project.

SI-Home study is a multidisciplinary working group of Siriraj Hospital to together implement appropriate public health interventions in response to COVID-19. These actions include active and passive surveillance, active case finding, contact tracing, preparation of resource, laboratory capacity to diagnose SARS-CoV-2 infection, quarantine facilities and services, etc. The emphasis is on efficient response with appropriate target populations.

**Siriraj Executive Committee**

Prasit Wattanapa^1^, Visit Vamvanij^2^, Chamaiporn Charoenkraikamol^3^, Daranee Piputtanakulchai^3^

^1^Dean, Faculty of Medicine Siriraj Hospital, Mahidol University; ^2^Director, Faculty of Medicine Siriraj Hospital, Mahidol University; ^3^Department of Nursing, Faculty of Medicine Siriraj Hospital, Mahidol University

**Medical Monitoring Team**

Pochamana Phisalprapa^1^, Naruemit Sayabovorn^1^, Weerachai Srivanichakorn^1^, Chaiwat Washirasaksiri^1^, Chonticha Auesomwang^1^, Tullaya Sitasuwan^1^, Thanet Chaisathaphol^1^, Rungsima Tinmanee^1^, Methee Chayakulkeeree^2^, Pakpoom Phoompoung^2^, Gornmigar, Sanpawitayakul^3^, Rungsima Wanitphakdeedecha^4^, Saipin Muangman^5^, Korapat Mayurasakorn^6^

^1^Division of Ambulatory Medicine, Department of Medicine, Faculty of Medicine Siriraj Hospital, Mahidol University; ^2^Division of Infectious Diseases and Tropical Medicine, Department of Medicine, Faculty of Medicine Siriraj Hospital, Mahidol University; ^3^Department of Pediatrics, Faculty of Medicine Siriraj Hospital, Mahidol University; ^4^Department of Dermatology, Faculty of Medicine Siriraj Hospital, Mahidol University; ^5^Department of Anesthesiology, Faculty of Medicine Siriraj Hospital, Mahidol University; ^6^Siriraj Population Health and Nutrition Research Group (SPHERE), Department of Research Group and Research Network, Siriraj Medical Research Center, Mahidol University

**Si-Home Management Team**

Sureeporn Pumeiam^1^ (coordinator), Bonggochpass Pinsawas^1^ (online coordinator), Pichanun Mongkolsucharitkul^1^ (online coordinator), Apinya Surawit^1^ (data analyzer), Tanyaporn Pongkunakorn^1^ (online coordinator), Sophida Suta^1^ (online coordinator), Thamonwan Manosan^1^ (online coordinator), Suphawan Ophakas^1^ (online coordinator), Nattvara Nirdnoy^1^ (online coordinator), Korapat Mayurasakorn^1^ (online supervisor), Suree Leemongkol(continuing care manager)^2^, Usanee Fongsri^2^, Pranom Sahunphun^2^, Panita Janthapadchothe^3^

^1^Siriraj Population Health and Nutrition Research Group (SPHERE), Department of Research Group and Research Network, Siriraj Medical Research Center, Mahidol University; ^2^Department of Nursing, Faculty of Medicine Siriraj Hospital, Mahidol University; ^3^Pharmacy Department, Faculty of Medicine Siriraj Hospital, Mahidol University

**SI-Home Operational Team**

Penrapee Phoosongtham^1^ (site manager), Wanwalee Kochasawas^1^ (nurse case manager), Phakamas Jitpun^1^ (charge nurse)

^1^Department of Nursing, Faculty of Medicine Siriraj Hospital, Mahidol University

**Study source recruitment and enrollment**

The study population was all COVID-19 patients who were treated in HI system of Siriraj Hospital. Participants included 2704 patients in the Delta pandemic (July to October 2021) and 2477 patients in the Omicron pandemic (January to March 2022). We monitored general demographic conditions, presence of comorbidities, onset symptoms of COVID-19, and vaccination history. Immunogenicity evaluation and neutralizing antibody against variants and post COVID-19 symptoms after recovery at 21-150 days post COVID-19 onset were investigated in a subset of 495 patients during the Delta pandemic.

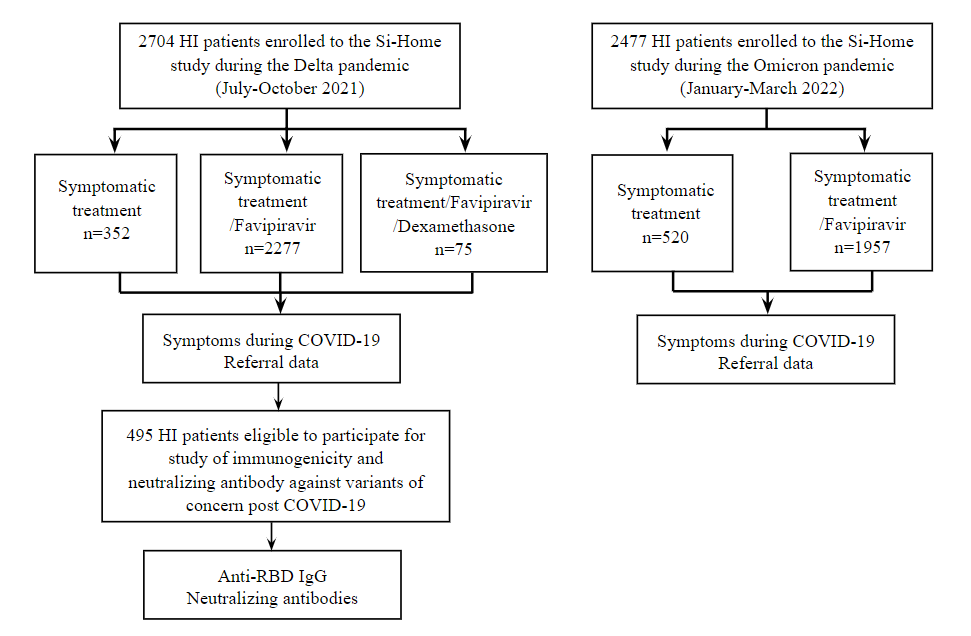

**Figure S1: Study source recruitment and enrollment**

**Methods S1: Study design, eligibility, inclusion and exclusion criteria**

**Study design**

This was a multidisciplinary, retrospective cohort study involving COVID-19 patients admitted at HI system and monitored by remote medical consultation in Bangkok, Thailand. A total of 11,897 patients with mild to moderate COVID-19 were isolated at home in Siriraj HI system during July 8, 2021 to May 11, 2022. The data collection reported here was performed between July 8, 2021 to March 15, 2022. The study population included 2704 and 2477 patients during the Delta (before November 2021) and Omicron (after January 12, 2022) pandemic who had been positive with SARS-CoV-2 as determined by a reverse transcriptase polymerase chain reaction (RT-PCR) testing. Someone who met inclusion criteria would be considered to have mild symptoms, or perhaps be asymptomatic, and would be referred to Siriraj-Home system (SI-Home) where medicine will be delivered by health personnel within 24 hours rather than being relegated to a field hospital or other potentially unpleasant arrangement. A subset of 495 patients during Delta pandemic (July-October 2021) were followed for serology testing between 21-120 days post COVID-19 onset. This study was approved by the Institutional Review Board of the Faculty of Medicine Siriraj Hospital, Mahidol University (SIRB protocol 463/2564; COA: Si 503/2021 and SIRB protocol 769/2564; COA: Si 833/2021).

**Inclusion criteria** for those considered as COVID-19 infection with mild-to-moderate conditions suitable for SI-Home

1. Age ≥12 years with any genders
2. Well-controlled chronic underlying diseases (e.g. dyslipidemia, diabetes mellitus, hypertension)
3. Body temperature: below 39 degrees Celsius during PCR testing day
4. Able to use online communication platforms
5. Oxygen saturation (more than a 95% oxygen saturation value)
6. No evidence of pneumonia on an admission date
7. All patients in HI system were subject to a maximum of 10-14 days home quarantine according to a governmental guideline^1^

**Criteria for referral**

1. Anyone with a fever that stays above 39 degrees Celsius for more than 24 hours
2. Anyone with less than a 94% oxygen saturation value
3. Any adult who is inhaling faster than 25 breaths per minute
4. Children who have difficulty breathing or experience rapid breathing, loss of appetite, drowsiness, as well as a fever over 39 degrees Celsius and an oxygen saturation value under 94%

**Testing and monitoring processes**

All patients in this study were admitted in HI system. During isolation in the HI, patients had their temperatures and respiratory symptoms checked ≥2 times each day, either by telemedicine with nurses or by using self-monitoring equipment and reported via an electronic diary. Online (24 hours/7 days) communication with medical staff was a standard management to monitor progression of the COVID-19 for medical management. Customized sets of medication would be provided for each patient for symptomatic treatment, such as antipyretics antitussives and cetirizine, which were prescribed by a medical staff via an online platform. Online medical staff included infectious disease physicians, infectious pediatricians, ambulatory physicians, psychiatrists, family physicians, obstetricians, registered nurses, volunteers, adverse drug reaction monitoring pharmacists, dietitians and the referral center staff. Breathing exercises with prone position was a recommended technique for patients with dyspnea and reduced oxygen saturation. Sit-to-stand tests for exercise-induced desaturation was one of the techniques to verify a so-called “happy hypoxia”.^2^ Patients who developed symptoms such as dyspnea, chest pain, or chest discomforts, which did not subside after recommended treatment were transferred back to the hospital.

The days of post symptom onset (PSO) was determined based on the difference between the date of the RT-PCR testing and the date of first reported symptoms coherent with COVID-19. For asymptomatic individuals, the day from the first positive SARS-CoV-2 PCR testing was used. The durations of illness onset to first hospital admission, to first Favipiravir treatment and to discharge up to 14 days were measured. Favipiravir was recommended anti-viral therapy especially in patients with high risk for COVID-19 pneumonia within 48 hours and was given for 5 days, but duration can be extended to as long as 14 days based on patient’s clinical severity. The dose was 1800 mg twice daily on the first day followed by 800 mg twice daily.^3^

**A subpopulation follow-up study**

A subset of 495 participants during the Delta pandemic provided general information, local and systemic symptoms, and a vaccination history. Vaccine history of participants, including type, number of doses, and date(s) of vaccination, as described in the COVID-19 Application Privacy Notice (“Morprom”), if available. Additional blood samples were collected via phlebotomy in ethylenediaminetetraacetic acid (EDTA) tubes and processed for plasma between 21-120 days post COVID-19 onset. All sera were obtained and stored at -80 °C within Siriraj Biobank, Department of Research, Siriraj Medical Research Center, Mahidol University. All data, including demographic, clinical, laboratory, and therapy details were collected from a database of HI system after July 2021. Participants who were vaccinated after being discharged from HI were excluded. Additional plasma samples were collected.

At the study visit, all patients were classified into one of different exposure groups based on vaccination status prior to COVID-19 infection, study antibody and PCR test. Groups were included (1) patients who had not had any vaccination, (2) patients who had a first vaccination <14 days before infection, (3) patients who had a first vaccination ≥14 days before infection, (4) patients who had a second vaccination <14 days before infection, (5) patients who had a second vaccination ≥14 days before infection, and on rare occasion had (6) patients who had a third vaccination <14 days before infection, and (7) patients who had a third vaccination ≥14 days before infection.

**SARS-CoV-2 PCR testing**

Our COVID-19 diagnostic assay was a probe-based qualitative RT-PCR probe. Allplex™ 2019-nCoV Assay (Seegene, Seoul, South Korea) was used for SARS-CoV2 detection. The targeted COVID-19 genes detected here included nucleocapsid (N), envelope (E) of Sarbecovirus and RNA-dependent RNA polymerase (RdRp) of COVID-19 according to the manufacturer’s instructions and described previously.^4^

**Detection of SARS-CoV-2 antibodies and surrogate virus neutralization test** **against SARS-CoV-2 strains**

One serological test was done during post-COVID-19. A subgroup of participants who received either symptomatic treatment vs. Favipiravir treatment vs. Favipiravir and Dexamethasone were randomly invited to test for anti-SARS-CoV-2 receptor binding domain IgG. A chemiluminescent microparticle immunoassay for qualitative detection of IgG against RBD of the SARS-CoV-2 spike protein (S1 subunit, No. 06S60) and SARS-CoV-2 nucleocapsid protein (SARS-CoV-2 IgG II Quant for use with ARCHITECT; Abbott Laboratories, Abbott Park, IL, USA; reference 06R8620) was undertaken.^7^ This assay linearly measures the level of antibody between 21.0-40,000.0 arbitrary unit (AU)/mL, which was converted later to WHO International Standard concentration as binding antibody unit per mL (BAU/mL) following the equation provided by the manufacturer (BAU/mL=0.142 x AU/mL). The level greater or equal to the cutoff value of 50 AU/mL or 7.1 BAU/mL was defined as seropositive. Briefly, plasma was pre-incubated with horseradish peroxidase conjugated-receptor binding domain protein (HRP-conjugated RBD protein). Subsequently, mixture was transferred to each well containing Streptavidin bound with Biotin conjugated angiotensin-converting enzyme 2 (ACE2). The plate was washed, substrate and stop solution were added. Finally, the optical density absorbance was measured using spectrophotometer at 450 nm. Inhibition rate was calculated through this formula:

$$Inhibition rate \left( \% \right)=\left( 1-\frac{{OD}_{450} of Sample}{{OD}_{450} of Negative control} \right) x 100.$$

Sample diluent was used as the negative control. White blood cell count, C-reactive protein, and D-dimer results were retrieved from electronic medical record from patients who were readmitted to the hospital.

**Outcomes and statistical analysis**

The characteristics of the patients at baseline were reported as counts and percentages or, for continuous variables, as medians with interquartile ranges. For the descriptive analysis, categorical variables were presented as absolute numbers and their relative frequencies. Continuous variables were summarized as mean and standard deviation if normally distributed or as the median and interquartile range (IQR) if not normally distributed. Comparisons of Ct values by vaccine exposure groups used quantile (median) regression adjusted for age and sex (Figure 1) in full text of this article. Patients who responded to the questionnaire were included in the analysis, and results are presented as percentages with means or medians and 95% confidence interval (CI). In univariable analysis, categorical variables were compared using the chi-square test and binomial logistic regression and presented with ORs, 95% CI and *p*-values. Multivariable analysis was performed by binary logistic regression for dichotomous outcome variables (Tables S4-5). Cox regression analysis was used to analyze the factors of the negative conversion time (NCT) of SARS-CoV-2 RNA (NCT is closely related to clinical manifestation and disease progression in COVID-19 patients. First, univariate analysis was performed, and the indicators with statistical significance were analyzed for Kaplan-Meier survival analysis. A Cox proportional hazard model was used for multivariate analysis (Table S6 and Figure S3). The differences in continuous and categorical data were tested using the Mann-Witney U test or t-test, Kruskal-Wallis H test or Analysis of variance (ANOVA) and chi-square test, respectively. The anti-SARS-CoV-2 RBD IgG concentration and neutralization antibodies were reported as geometric mean titers (GMT) with 95% CI. Unpaired t test, and ANOVA were used to compare GMT within group, between groups, and across groups using GraphPad Prism 9 version 9.2.0 (GraphPad Software, CA, USA), respectively (Tables S7-10 and Figure S4). All analyses were conducted in STATA version 17 (Stata Corp, LP, College Station, TX, USA), and graphs were produced in GraphPad Prism 9 (GraphPad Software, CA, USA).

**Trend of SARS-CoV-2 variants circulating in Thailand**

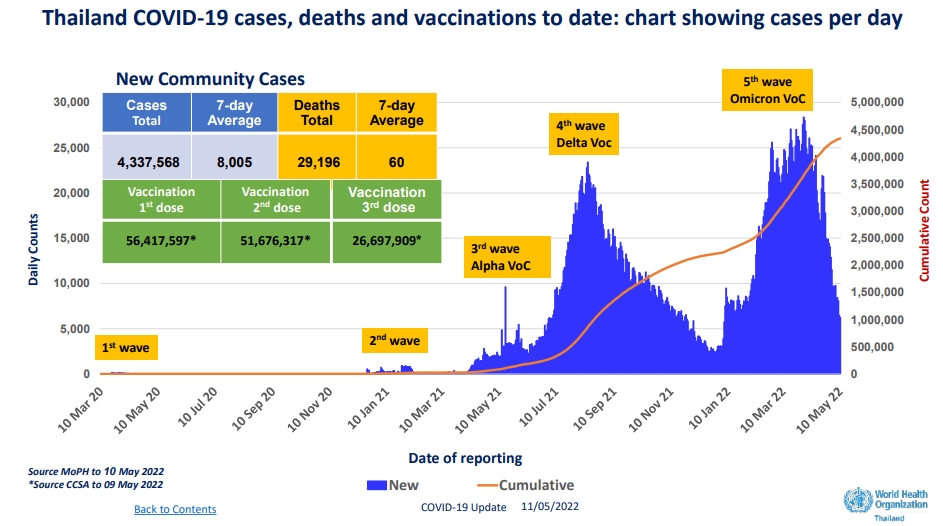

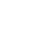

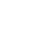

**Figure S2: Trend of SARS-CoV-2 variants circulating in Thailand, deaths and vaccination status during analysis period^5^**

The drop in new cases over the last seven days decrease by 35% compared to the previous week. Most cases continue to be monitored in satellite facilities (hospitels, community isolation and home isolation). The average number of COVID cases occupying hospital beds per day over the past week decreases by 21% compared with the week prior. vaccination rates in Thailand continue to significantly reduce levels of severe illness and deaths caused by circulating Omicron variants. High vaccination rates also help ameliorate the transmission of COVID-19.

| **Table S1: General comorbidity questionnaire** | | | | | | | | |
| --- | --- | --- | --- | --- | --- | --- | --- | --- |
| **Questions** | | | **Answer 1** | | **Answer 2** | | **Answer 3** | |
| **General information** | | | | | | | | |
| 1. | | Age | __________ years | | | | | |
| 2. | | Gender | Male | | Female | |  | |
| 3. | | Pregnancy status | Yes (___ weeks) | | No | |  | |
| 4 | | Career | ___________ | | | | | |
| 5. | | Do you have any comorbidities? (chronic upper respiratory tract disease, cardiovascular disease, chronic kidney disease, cerebrovascular disease, obesity, cancer, uncontrolled diabetes) | Yes | | No | |  | |
| 6. | | Did you experience COVID-19 infection in the previous 3 months? | Yes | | No | | Not sure | |
| 7. | | Did you have a history of COVID-19 reinfection? | Yes | | No | | Not sure | |
| **Evaluation of previous health status** | | | | | | | | |
| 8. | | Did you experience significant weight loss? | Yes | | No | | Not sure | |
| 9. | | Did you experience anorexia and nausea? | Yes | | No | | Not sure | |
| 10. | | Did you experience erectile dysfunction? | Yes | | No | | Not sure | |
| 11. | | Did you experience menstrual changes? | Yes | | No | | Not sure | |
| 12. | | Did you experience being unable to control movement? | Yes | | No | | Not sure | |
| 13. | | Did you experience feeling of a numb body or face? | Yes | | No | | Not sure | |
| 14. | | Did you experience stability issues? | Yes | | No | | Not sure | |
| 15. | | Did you experience dysphagia problems? | Yes | | No | | Not sure | |
| 16. | | Did you experience vision problems? | Yes | | No | | Not sure | |
| 17. | | Did you experience communication problems? | Yes | | No | | Not sure | |
| 18. | | Did you experience sleep problems? | Yes | | No | | Not sure | |
| 19. | | Did you experience urinary tract problems? | Yes | | No | | Not sure | |
| **Evaluation of adverse signs and symptoms in the past 7 days** | | | | | | | | |
| 20. | | Did you experience fever? | Yes | | No | | Not sure | |
| 21. | | Did you experience headache? | Yes | | No | | Not sure | |
| 22. | | Did you experience cough? | Yes | | No | | Not sure | |
| 23. | | Did you experience sore throat? | Yes | | No | | Not sure | |
| 24. | | Did you experience running nose? | Yes | | No | | Not sure | |
| 25. | | Did you experience shortness of breath or difficulty breathing? | Yes | | No | | Not sure | |
| 26. | | Did you experience chest pain? | Yes | | No | | Not sure | |
| 27. | | Did you experience myalgia or weakness? | Yes | | No | | Not sure | |
| 28. | | Did you experience joint pain? | Yes | | No | | Not sure | |
| 29. | | Did you experience fatigue? | Yes | | No | | Not sure | |
| 30. | | Did you experience leg/arm numbness? | Yes | | No | | Not sure | |
| 31. | | Did you experience confusion or stupor? | Yes | | No | | Not sure | |
| 32. | | Did you experience any seizure? | Yes | | No | | Not sure | |
| 33. | | Did you experience abdominal pain or diarrhea? | Yes | | No | | Not sure | |
| 34. | | Did you experience nausea, diarrhea or vomiting? | Yes | | No | | Not sure | |
| 35. | | Did you experience conjunctivitis or blurred vision? | Yes | | No | | Not sure | |
| 36. | | Did you experience rash? | Yes | | No | | Not sure | |
| **COVID-19 vaccination histories** | | | | | | | | |
| 37. | | Did you get any COVID-19 vaccines before? | Yes | | No | | Not sure | |
| 38. | | How many shots of COVID-19 vaccine? | ___________ shot(s) | | | | | |
| 39. | | Types of COVID-19 vaccination (Sinovac, AstraZeneca, Sinopharm, Pfizer, Moderna, Johnson& Johnson, Novavax, Sputnik V) | ____________ | | | | | |
| 40. | | Did you experience an allergic reaction? If so, please explain. | Yes | | No | | Not sure | |
| 41. | | Did you experience other symptoms after receiving vaccines? If so, please explain. | Yes | | No | | Not sure | |
| **Symptoms during COVID-19 care in HI system** | | |  | |  | |  | |
| 1. | | Did you experience fever? | Yes | | No | | Not sure | |
| 2. | | Did you experience cough? | Yes | | No | | Not sure | |
| 3. | | Did you experience sore throat? | Yes | | No | | Not sure | |
| 4. | | Did you experience Rhinorrhea? | Yes | | No | | Not sure | |
| 5. | | Did you experience Productive sputum? | Yes | | No | | Not sure | |
| 6. | | Did you experience Loss of taste? | Yes | | No | | Not sure | |
| 7. | | Did you experience Loss of smell? | Yes | | No | | Not sure | |
| 8. | | Did you experience Dyspnea? | Yes | | No | | Not sure | |
| 9. | | Did you experience Myalgia? | Yes | | No | | Not sure | |
| 10. | | Did you experience Diarrhea? | Yes | | No | | Not sure | |
| 11. | | Did you experience Nausea/vomiting? | Yes | | No | | Not sure | |
| 12. | | Did you experience Others? | Yes | | No | | Not sure | |
| **Questions of chalder fatigue scale*** | | | **less than**  **usual**  **(0)** | **no more**  **than usual**  **(1)** | | **more than**  **usual**  **(2)** | | **much more**  **than usual**  **(3)** |
| 1. | Do you have problems with tiredness? | |  |  | |  | |  |
| 2. | Do you feel sleepy or drowsy? | |  |  | |  | |  |
| 3. | Do you lack energy? | |  |  | |  | |  |
| 4. | Do you have less strength in your muscles? | |  |  | |  | |  |
| 5. | Do you feel weak? | |  |  | |  | |  |
| 6. | Do you have difficulties concentrating? | |  |  | |  | |  |
| 7. | Do you need to rest more? | |  |  | |  | |  |
| 8. | Do you have problems starting things? | |  |  | |  | |  |
| ­­­9. | How is your memory? | |  |  | |  | |  |

*This scale can be scored “bimodally” with columns representing 0 & 1 and a range from 0 to 9 with a total of 4 or more qualifying for “caseness”. Alternatively, it can be stored in “Likert” style 0, 1, 2 & 3 with a range from 0 to 27.

| **Table S2: Characteristics of participants at enrollment (Jul-Oct 2021 (Delta)) by age** | | | | | | | | | | | | | | | | | | | |
| --- | --- | --- | --- | --- | --- | --- | --- | --- | --- | --- | --- | --- | --- | --- | --- | --- | --- | --- | --- |
| **Characteristics** | | | | **Jul-Oct 2021 (Delta)** | | | | | | | | | | | | | | | |
|  |  |  |  | **All patients** | | **0-11 years** | | **12-18 years** | | **19-30 years** | | **31-45 years** | | | **46-60 years** | | | **>60 years** | |
|  |  |  |  | **n** | **%** | **n** | **%** | **n** | **%** | **n** | **%** | **n** | | **%** | **n** | **%** | | **n** | **%** |
|  |  |  |  | **n=2704** | | **n=448** | | **n=227** | | **n=560** | | **n=677** | | | **n=513** | | | **n=279** | |
| Age, years, mean (SD) | | | | 33.8 | (19.6) | 5.2 | (3.4) | 15.2 | (2.1) | 24.7 | (3.3) | 38.3 | (4.4) | | 52.8 | | (4.4) | 67.0 | (5.6) |
| Gender | | | |  |  |  |  |  |  |  |  |  |  | |  | |  |  |  |
|  | male | | | 1253 | 46.3 | 221 | 49.3 | 115 | 50.7 | 266 | 47.5 | 318 | 47.0 | | 225 | | 43.9 | 108 | 38.7 |
|  | female | | | 1451 | 53.7 | 227 | 50.7 | 112 | 49.3 | 294 | 52.5 | 359 | 53.0 | | 288 | | 56.1 | 171 | 61.3 |
| Body weight, kg, median (IQR) | | | | 57.5 | (22.0) | 23.2 | (13.6) | 56.4 | (18.6) | 65.9 | (17.3) | 67.1 | (14.8) | | 65.9 | | (13.6) | 62.6 | (11.7) |
| ***Presence of comorbidities*** | | | | | | | | | | | | | | | | | | | |
|  | Diabetes mellitus | | | 162 | 6.0 | 0 | 0.0 | 0 | 0.0 | 3 | 0.5 | 19 | 2.8 | | 67 | | 13.1 | 73 | 26.2 |
|  | Hypertension | | | 313 | 11.6 | 0 | 0.0 | 0 | 0.0 | 9 | 1.6 | 48 | 7.1 | | 118 | | 23.0 | 138 | 49.5 |
|  | Dyslipidemia | | | 123 | 4.6 | 0 | 0.0 | 0 | 0.0 | 1 | 0.2 | 18 | 2.7 | | 46 | | 9.0 | 58 | 20.8 |
|  | Obesity | | | 23 | 0.9 | 2 | 0.5 | 4 | 1.8 | 4 | 0.7 | 8 | 1.2 | | 3 | | 0.6 | 2 | 0.7 |
|  | Malignancy | | | 21 | 0.8 | 0 | 0.0 | 0 | 0.0 | 3 | 0.5 | 4 | 0.6 | | 6 | | 1.2 | 8 | 2.9 |
|  | Neurologic disease | | | 7 | 0.3 | 0 | 0.0 | 0 | 0.0 | 2 | 0.4 | 1 | 0.2 | | 4 | | 0.8 | 0 | 0.0 |
|  | Heart disease | | | 33 | 1.2 | 2 | 0.5 | 1 | 0.4 | 1 | 0.2 | 5 | 0.7 | | 13 | | 2.5 | 11 | 3.9 |
|  | Lung disease | | | 51 | 1.9 | 1 | 0.2 | 0 | 0.0 | 2 | 0.4 | 9 | 1.3 | | 19 | | 3.7 | 20 | 7.2 |
|  | Kidney disease | | | 14 | 0.5 | 0 | 0.0 | 0 | 0.0 | 2 | 0.4 | 3 | 0.4 | | 3 | | 0.6 | 6 | 2.2 |
|  | Others | | | 336 | 12.4 | 43 | 9.6 | 22 | 9.7 | 57 | 10.2 | 90 | 13.3 | | 84 | | 16.4 | 40 | 14.3 |
|  | Dead after referral | | | 5 | 0.2 | 0 | 0.0 | 0 | 0.0 | 0 | 0.0 | 0 | 0.0 | | 3 | | 0.6 | 2 | 0.7 |
| ***Presenting symptoms*** | | | | | | | | | | | | | | | | | | | |
|  | Asymptomatic infection | | | 390 | 14.4 | 99 | 22.1 | 32 | 14.1 | 42 | 7.5 | 87 | 12.9 | | 79 | | 15.4 | 51 | 18.3 |
|  | Fever/history of fever | | | 1250 | 46.2 | 262 | 58.5 | 96 | 42.3 | 261 | 46.6 | 305 | 45.1 | | 208 | | 40.6 | 118 | 42.3 |
|  | BT(°C), median (IQR) | | | 36.6 | (0.7) | 36.3 | (1.0) | 36.3 | (0.6) | 36.7 | (0.5) | 36.7 | (0.5) | | 36.7 | | (0.5) | 36.7 | (0.8) |
|  |  | <37.5 | | 2474 | 95.2 | 387 | 93.3 | 203 | 96.2 | 514 | 95.0 | 632 | 96.1 | | 476 | | 94.8 | 262 | 96.0 |
|  |  | 37.5-38.0 | | 120 | 4.6 | 26 | 6.3 | 8 | 3.8 | 27 | 5.0 | 26 | 4.0 | | 26 | | 5.2 | 7 | 2.6 |
|  |  | >38.0 | | 6 | 0.2 | 2 | 0.5 | 0 | 0.0 | 0 | 0.0 | 0 | 0.0 | | 0 | | 0.0 | 4 | 1.5 |
|  | Cough | | | 1642 | 60.7 | 212 | 47.3 | 128 | 56.4 | 354 | 63.2 | 438 | 64.7 | | 332 | | 64.7 | 178 | 63.8 |
|  | Sore throat | | | 1038 | 38.4 | 63 | 14.1 | 82 | 36.1 | 286 | 51.1 | 303 | 44.8 | | 208 | | 40.6 | 96 | 34.4 |
|  | Rhinorrhea | | | 419 | 15.5 | 56 | 12.5 | 42 | 18.5 | 109 | 19.5 | 120 | 17.7 | | 55 | | 10.7 | 37 | 13.3 |
|  | Productive sputum | | | 537 | 19.9 | 40 | 8.9 | 45 | 19.8 | 140 | 25.0 | 170 | 25.1 | | 88 | | 17.2 | 54 | 19.4 |
|  | Loss of taste | | | 312 | 11.5 | 7 | 1.6 | 35 | 15.4 | 110 | 19.6 | 99 | 14.6 | | 46 | | 9.0 | 15 | 5.4 |
|  | Loss of smell | | | 821 | 30.4 | 21 | 4.7 | 88 | 38.8 | 266 | 47.5 | 263 | 38.9 | | 145 | | 28.3 | 38 | 13.6 |
|  | Dyspnea | | | 305 | 11.3 | 6 | 1.3 | 18 | 7.9 | 76 | 13.6 | 88 | 13.0 | | 73 | | 14.2 | 44 | 15.8 |
|  | Myalgia (muscle aches) | | | 282 | 10.4 | 5 | 1.1 | 22 | 9.7 | 70 | 12.5 | 108 | 16.0 | | 47 | | 9.2 | 30 | 10.8 |
|  | Diarrhea | | | 126 | 4.7 | 11 | 2.5 | 16 | 7.1 | 40 | 7.1 | 39 | 5.8 | | 11 | | 2.1 | 9 | 3.2 |
|  | Nausea/vomiting | | | 59 | 2.2 | 4 | 0.9 | 3 | 1.3 | 12 | 2.1 | 24 | 3.6 | | 8 | | 1.6 | 8 | 2.9 |
|  | Others | | | 1549 | 57.3 | 218 | 48.7 | 126 | 55.5 | 351 | 62.7 | 405 | 59.8 | | 285 | | 55.6 | 164 | 58.8 |
| ***Clinical features at the time of admission*** | | | | | | | | | | | | | | | | | | | |
|  | Time from symptom onset to PCR diagnosis, days, median (IQR) | | | 1.9 | (1.6) | 2.2 | (2.4) | 1.9 | (1.6) | 2.1 | (1.5) | 1.9 | (1.4) | | 1.6 | | (1.1) | 1.8 | (1.6) |
|  | Time from symptom onset to admission, days, median (IQR) | | | 5.1 | (2.4) | 4.8 | (2.6) | 5.4 | (2.6) | 5.1 | (2.2) | 5.1 | (2.3) | | 5.0 | | (2.3) | 5.0 | (2.3) |
| ***Cycle Threshold, median (IQR)*** | | | |  |  |  |  |  |  |  |  |  |  | |  | |  |  |  |
|  | Nucleocapsid (N) | | | 21.0 | (7.8) | 20.1 | (7.9) | 21.7 | (8.9) | 21.8 | (7.4) | 21.1 | (8.9) | | 21.0 | | (7.3) | 20.0 | (7.2) |
|  |  | | <20 | 1118 | 43.2 | 219 | 49.7 | 81 | 36.0 | 204 | 38.1 | 267 | 42.3 | | 213 | | 43.4 | 134 | 50.4 |
|  |  | | 20-30 | 1218 | 47.0 | 174 | 39.5 | 117 | 52.0 | 284 | 53.0 | 301 | 47.6 | | 230 | | 46.8 | 112 | 42.1 |
|  |  | | >30 | 255 | 9.8 | 48 | 10.9 | 27 | 12.0 | 48 | 9.0 | 64 | 10.1 | | 48 | | 9.8 | 20 | 7.5 |
|  | Envelope (E) | | | 17.5 | (8.4) | 16.9 | (8.4) | 18.4 | (9.2) | 18.3 | (7.9) | 17.7 | (9.6) | | 17.5 | | (7.7) | 16.3 | (7.9) |
|  |  | | <20 | 1713 | 64.7 | 292 | 66.7 | 135 | 60.5 | 344 | 62.4 | 415 | 62.2 | | 341 | | 67.5 | 186 | 70.2 |
|  |  | | 20-30 | 838 | 31.6 | 134 | 30.6 | 81 | 36.3 | 187 | 33.9 | 225 | 33.7 | | 140 | | 27.7 | 71 | 26.8 |
|  |  | | >30 | 98 | 3.7 | 12 | 2.7 | 7 | 3.1 | 20 | 3.6 | 27 | 4.1 | | 24 | | 4.8 | 8 | 3.0 |
| RNA-dependent RNA polymerase (RdRp) | | | | 22.3 | (8.2) | 21.8 | (7.9) | 23.4 | (9.2) | 23.2 | (7.5) | 22.3 | (9.1) | | 22.1 | | (7.3) | 21.2 | (7.6) |
| Immunosuppression, median (IQR) | | | | 1060.6 | (1070.1) | - | - | 762 | (730.5) | 1794.9 | (1667.8) | 805.5 | (745.7) | | 1606.9 | | (1481.9) | 1224.3 | (1192.6) |
| Neutralizing antibody titers, median (IQR) | | | | 96.8 | (50.9) | - | - | 82.2 | (74.7) | 85.2 | (83.4) | 91.8 | (61.7) | | 94.3 | | (36.1) | 90.9 | (30.0) |

Physical symptoms information and laboratory analysis results of COVID-19 patients in the Delta wave by age range. Data are presented as mean (SD), median (IQR), and range at *p*<0.05 indicates statistical significance. SD, standard deviation; IQR, interquartile range; BT, body temperature; °C, degree Celsius, SARS-CoV-2 spike protein antibody titers, log_10_ transformed. Neutralizing antibody titers, log_10_ transformed.

| **Table S3: Characteristics of participants at enrollment (Jan-Mar 2022 (Omicron)) by age** | | | | | | | | | | | | | | | | | | | | | | | | | | | | | | | | | | | | | | | | | | | | | | | | | | | | | | |
| --- | --- | --- | --- | --- | --- | --- | --- | --- | --- | --- | --- | --- | --- | --- | --- | --- | --- | --- | --- | --- | --- | --- | --- | --- | --- | --- | --- | --- | --- | --- | --- | --- | --- | --- | --- | --- | --- | --- | --- | --- | --- | --- | --- | --- | --- | --- | --- | --- | --- | --- | --- | --- | --- | --- |
| **Characteristics** | | | | | | **Jan-Mar 2022 (Omicron)** | | | | | | | | | | | | | | | | | | | | | | | | | | | | | | | | | | | | | | | | | | | | | | | | |
|  |  |  |  |  |  | **All patients** | | | | | | | | | **0-11 years** | | | | | | | **12-18 years** | | | | | | **19-30 years** | | | | | | | | **31-45 years** | | | | | | | | **46-60 years** | | | | | | | **>60 years** | | | |
|  |  |  |  |  |  | **n** | | | **%** | | | | | | **n** | | | **%** | | | | **n** | | | | **%** | | **n** | | | | | **%** | | | **n** | | | | **%** | | | | **n** | **%** | | | | | | **n** | | **%** | |
|  |  |  |  |  |  | **n=2447** | | | | | | | | | **n=516** | | | | | | | **n=234** | | | | | | **n=544** | | | | | | | | **n=518** | | | | | | | | **n=360** | | | | | | | **n=242** | | | |
| Age, years, mean (SD) | | | | | | 31.3 | | | (20.3) | | | | | | 5.6 | | | (3.5) | | | | 14.9 | | | | (1.8) | | 24.8 | | | | | (3.3) | | | 37.8 | | | | (4.4) | | | | 52.4 | (4.4) | | | | | | 69.8 | | (7.4) | |
| Gender | | | | | | | | | | | | | | | | | | | | | | | | | | | | | | | | | | | | | | | | | | | | | | | | | | | | | | |
|  | male | | | | | 1031 | | 41.6 | | | | 269 | | | | | | | 52.1 | 117 | | | | 50.0 | | | | | 203 | | | | 37.3 | | | | 202 | | 34.8 | | | 137 | | | | | 38.1 | | | | 103 | | | 42.6 |
|  | female | | | | | 1446 | | 58.4 | | | | 247 | | | | | | | 47.9 | 117 | | | | 50.0 | | | | | 341 | | | | 62.7 | | | | 379 | | 65.2 | | | 223 | | | | | 61.9 | | | | 139 | | | 57.4 |
|  | Body weight, kg, median (IQR) | | | | | 55.8 | | (22.4) | | | | 24.7 | | | | | | | (16.1) | 59.0 | | | | (18.4) | | | | | 62.4 | | | | (16.0) | | | | 66.7 | | (16.1) | | | 65.9 | | | | | (12.8) | | | | 61.1 | | | (13.6) |
| ***Presence of comorbidities*** | | | | | | | | | | | | | | | | | | | | | | | | | | | | | | | | | | | | | | | | | | | | | | | | | | | | | | |
|  | Diabetes mellitus | | | | | | 150 | 6.1 | | | | | | 0 | | 0.0 | | | | | 1 | | | | 0.4 | | 2 | | | | 0.4 | | | | 25 | | | | 4.8 | | | 49 | | | | | 13.6 | | | | 73 | | 30.2 | |
|  | Hypertension | | | | | | 513 | 20.7 | | | | | | 32 | | 6.2 | | | | | 25 | | | | 10.7 | | 63 | | | | 11.6 | | | | 108 | | | | 20.9 | | | 140 | | | | | 38.9 | | | | 145 | | 59.9 | |
|  | Dyslipidemia | | | | | | 108 | 4.4 | | | | | | 0 | | 0.0 | | | | | 0 | | | | 0.0 | | 0 | | | | 0.0 | | | | 19 | | | | 3.7 | | | 35 | | | | | 9.7 | | | | 54 | | 22.3 | |
|  | Obesity | | | | | | 41 | 1.7 | | | | | | 5 | | 1.0 | | | | | 6 | | | | 2.6 | | 5 | | | | 0.9 | | | | 13 | | | | 2.5 | | | 11 | | | | | 3.1 | | | | 1 | | 0.4 | |
|  | Malignancy | | | | | | 25 | 1.0 | | | | | | 0 | | 0.0 | | | | | 0 | | | | 0.0 | | 0 | | | | 0.0 | | | | 0 | | | | 0.0 | | | 10 | | | | | 2.8 | | | | 15 | | 6.2 | |
|  | Neurologic disease | | | | | | 271 | 10.9 | | | | | | 12 | | 2.3 | | | | | 20 | | | | 8.6 | | 95 | | | | 17.5 | | | | 94 | | | | 18.2 | | | 37 | | | | | 10.3 | | | | 13 | | 5.4 | |
|  | Heart disease | | | | | | 38 | 1.5 | | | | | | 3 | | 0.6 | | | | | 0 | | | | 0.0 | | 1 | | | | 0.2 | | | | 8 | | | | 1.5 | | | 10 | | | | | 2.8 | | | | 16 | | 6.6 | |
|  | Lung disease | | | | | | na |  | | | | | | na | |  | | | | | na | | | |  | | na | | | |  | | | | na | | | |  | | | na | | | | |  | | | | na | |  | |
|  | Kidney disease | | | | | | 10 | 0.4 | | | | | | 1 | | 0.2 | | | | | 0 | | | | 0.0 | | 0 | | | | 0.0 | | | | 1 | | | | 0.2 | | | 6 | | | | | 1.7 | | | | 2 | | 0.8 | |
|  | Others | | | | | | 753 | 30.4 | | | | | | 75 | | 14.5 | | | | | 44 | | | | 18.8 | | 90 | | | | 16.5 | | | | 169 | | | | 32.6 | | | 191 | | | | | 53.1 | | | | 184 | | 76.0 | |
|  | Dead after referral | | | | | | 2 | 0.1 | | | | | | 0 | | 0.0 | | | | | 0 | | | | 0.0 | | 0 | | | | 0.0 | | | | 0 | | | | 0.0 | | | 0 | | | | | 0.0 | | | | 2 | | 0.8 | |
| ***Presenting symptoms*** | | | | | | | | | | | | | | | | | | | | | | | | | | | | | | | | | | | | | | | | | | | | | | | | | | | | | | |
|  | Asymptomatic infection | | | | | | 969 | | | 39.1 | | 266 | | | | 51.6 | | | | 99 | | | | | | 42.3 | | 167 | | | | 30.7 | | | 183 | | | | | 35.3 | | | 142 | | | | | 39.4 | | | 112 | | 46.3 | |
|  | Fever/history of fever | | | | | | 267 | | | 10.8 | | 156 | | | | 30.2 | | | | 35 | | | | | | 15.0 | | 31 | | | | 5.7 | | | 19 | | | | | 3.7 | | | 12 | | | | | 3.3 | | | 14 | | 5.8 | |
|  | BT (°C), median (IQR) | | | | | | 36.8 | | | (0.5) | | 36.9 | | | | (1.3) | | | | 36.8 | | | | | | (0.8) | | 36.8 | | | | (0.4) | | | 36.7 | | | | | (0.4) | | | 36.7 | | | | | (0.3) | | | 36.8 | | (0.4) | |
|  | |  | | <37.5 | | | 2107 | | | | 88.2 | | | 317 | | 64.8 | | | | | | | 172 | | | 71.1 | | 509 | | | | | 88.2 | | | | 557 | | | 90.9 | | | 342 | | | | | 87.5 | | | 210 | | 80.5 | |
|  | |  | | 37.5-38.0 | | | 244 | | | | 10.2 | | | 135 | | 27.6 | | | | | | | 33 | | | 13.6 | | 31 | | | | | 5.4 | | | | 19 | | | 3.1 | | | 12 | | | | | 3.1 | | | 14 | | 5.4 | |
|  | |  | | >38.0 | | | 23 | | | | 1.0 | | | 21 | | 4.3 | | | | | | | 2 | | | 0.8 | | 0 | | | | | 0.0 | | | | 0 | | | 0.0 | | | 0 | | | | | 0.0 | | | 0 | | 0.0 | |
|  | | Cough | | | | | 1181 | | | | 47.7 | | | 163 | | 31.6 | | | | | | | 107 | | | 45.7 | | 319 | | | | | 58.6 | | | | 322 | | | 62.2 | | | 168 | | | | | 46.7 | | | 102 | | 42.2 | |
|  | | Sore throat | | | | | 1181 | | | | 47.7 | | | 163 | | 31.6 | | | | | | | 107 | | | 45.7 | | 319 | | | | | 58.6 | | | | 322 | | | 62.2 | | | 168 | | | | | 46.7 | | | 102 | | 42.2 | |
|  | | Rhinorrhea | | | | | 626 | | | | 25.3 | | | 85 | | 16.5 | | | | | | | 56 | | | 23.9 | | 165 | | | | | 30.3 | | | | 181 | | | 34.9 | | | 89 | | | | | 24.7 | | | 50 | | 20.7 | |
|  | | Productive sputum | | | | | na | | | |  | | | na | |  | | | | | | | na | | |  | | na | | | | |  | | | | na | | |  | | | na | | | | |  | | | na | |  | |
|  | | Loss of taste | | | | | 43 | | | | 1.7 | | | 0 | | 0.0 | | | | | | | 2 | | | 0.9 | | 17 | | | | | 3.1 | | | | 19 | | | 3.7 | | | 5 | | | | | 1.4 | | | 0 | | 0.0 | |
|  | | Loss of smell | | | | | 43 | | | | 1.7 | | | 0 | | 0.0 | | | | | | | 2 | | | 0.9 | | 17 | | | | | 3.1 | | | | 19 | | | 3.7 | | | 5 | | | | | 1.4 | | | 0 | | 0.0 | |
|  | | Dyspnea | | | | | 24 | | | | 1.0 | | | 3 | | 0.6 | | | | | | | 2 | | | 0.9 | | 5 | | | | | 0.9 | | | | 7 | | | 1.4 | | | 4 | | | | | 1.1 | | | 3 | | 1.2 | |
|  | | Myalgia (muscle aches) | | | | | 206 | | | | 8.3 | | | 10 | | 1.9 | | | | | | | 12 | | | 5.1 | | 59 | | | | | 10.9 | | | | 68 | | | 13.1 | | | 46 | | | | | 12.8 | | | 11 | | 4.6 | |
|  | | Diarrhea | | | | | 63 | | | | 2.5 | | | 13 | | 2.5 | | | | | | | 2 | | | 0.9 | | 17 | | | | | 3.1 | | | | 19 | | | 3.7 | | | 8 | | | | | 2.2 | | | 4 | | 1.7 | |
|  | | Nausea/vomiting | | | | | 37 | | | | 1.5 | | | 12 | | 2.3 | | | | | | | 3 | | | 1.3 | | 6 | | | | | 1.1 | | | | 11 | | | 2.1 | | | 4 | | | | | 1.1 | | | 1 | | 0.4 | |
|  | | Others | | | | | 191 | | | | 7.7 | | | 40 | | 7.8 | | | | | | | 16 | | | 6.8 | | 32 | | | | | 5.9 | | | | 51 | | | 9.9 | | | 29 | | | | | 8.1 | | | 23 | | 9.5 | |
| ***Clinical features at the time of admission*** | | | | | | | | | | | | | | | | | | | | | | | | | | | | | | | | | | | | | | | | | | | | | | | | | | | | | | |
|  | | | Time from symptom onset to PCR diagnosis, days, median (IQR) | | | 2.0 | | | | | (1.1) | | 2.2 | | | | (1.2) | | | | | 2.0 | | | | (1.1) | | 2.0 | | (1.0) | | | | 1.9 | | | | (1.0) | | | 2.0 | | | | | (1.0) | | | 1.8 | | | (0.9) | | |
|  | | | Time from symptom onset to admission, days, median (IQR) | | | 2.8 | | | | | (1.6) | | 2.3 | | | | (1.5) | | | | | 2.7 | | | | (1.6) | | 2.9 | | (1.5) | | | | 2.9 | | | | (1.6) | | | 3.0 | | | | | (1.6) | | | 3.0 | | | (1.6) | | |
| ***Cycle Threshold, median (IQR)*** | | | | | | | | | | | | | | | | | | | | | | | | | | | | | | | | | | | | | | | | | | | | | | | | | | | | | | |
|  | | | Nucleocapsid (N) | | | 19.0 | | | | | (5.7) | | 18.8 | | | | (4.7) | | | | | 19.3 | | | | (6.3) | | 20.5 | | (6.1) | | | | 20.4 | | | | (6.0) | | | 20.1 | | | | | (5.3) | | | | 20.6 | | (6.0) | | |
|  | | |  | | <20 | 1162 | | | | | 53.9 | | 280 | | | | 64.2 | | | | | 112 | | | | 54.3 | | 233 | | 46.5 | | | | 258 | | | | 47.7 | | | 172 | | | | | 50.9 | | | | 107 | | 45.8 | | |
|  | | |  | | 20-30 | 974 | | | | | 45.2 | | 137 | | | | 31.4 | | | | | 75 | | | | 36.4 | | 248 | | 49.5 | | | | 262 | | | | 48.5 | | | 146 | | | | | 43.2 | | | | 106 | | 45.4 | | |
|  | | |  | | >30 | 216 | | | | | 10.0 | | 21 | | | | 4.8 | | | | | 14 | | | | 6.8 | | 59 | | 11.8 | | | | 59 | | | | 10.9 | | | 40 | | | | | 11.8 | | | | 23 | | 9.8 | | |
|  | | | Envelope (E) | | | 18.0 | | | | | (5.4) | | 17.2 | | | | (4.3) | | | | | 17.5 | | | | (5.9) | | 18.4 | | (6.3) | | | | 18.5 | | | | (5.7) | | | 18.1 | | | | | (5.6) | | | | 18.4 | | (6.0) | | |
|  | | |  | | <20 | 1578 | | | | | 67.0 | | 340 | | | | 77.6 | | | | | 137 | | | | 68.2 | | 340 | | 63.0 | | | | 371 | | | | 64.1 | | | 235 | | | | | 65.6 | | | | 155 | | 65.1 | | |
|  | | |  | | 20-30 | 617 | | | | | 26.2 | | 83 | | | | 19.0 | | | | | 54 | | | | 26.9 | | 155 | | 28.7 | | | | 166 | | | | 28.7 | | | 95 | | | | | 26.5 | | | | 64 | | 26.9 | | |
|  | | |  | | >30 | 159 | | | | | 6.8 | | 15 | | | | 3.4 | | | | | 10 | | | | 5.0 | | 45 | | 8.3 | | | | 42 | | | | 7.3 | | | 28 | | | | | 7.8 | | | | 19 | | 8.0 | | |
|  | | | RNA-dependent RNA polymerase (RdRp) | | | 19.3 | | | | | (5.4) | | 18.6 | | | | (4.4) | | | | | 19.0 | | | | (5.6) | | 19.7 | | (5.9) | | | | 19.7 | | | | (5.8) | | | 19.3 | | | | | (5.4) | | | | 19.5 | | (5.5) | | |

Physical symptoms information and laboratory analysis results of COVID-19 patients in the Omicron wave by age range. Data are presented as mean (SD), median (IQR), and range at *p*<0.05 indicates statistical significance. SD, standard deviation; IQR, interquartile range; BT, body temperature; °C, degree Celsius, SARS-CoV-2 spike protein antibody titers, log_10_ transformed. Neutralizing antibody titers, log_10_ transformed.

| **Table S4: Characteristics of the first positive symptoms between the delta and omicron pandemics for people aged >18 years** | | | | | | | | | | | | |
| --- | --- | --- | --- | --- | --- | --- | --- | --- | --- | --- | --- | --- |
| **vaccination** | **Jul - Oct 2021 (Delta) n=2029** | | | | | | **Jan - Mar 2022 (omicron) n=1727** | | | | | |
|  | **Total** | **Any symptoms** | | **Cough, fever, anosmia, ageusia** | | ***p-value*** | **Total** | **Any symptoms** | | **Cough, fever, anosmia, ageusia** | | ***p-value*** |
|  |  | **n** | **%** | **n** | **%** |  |  | **n** | **%** | **n** | **%** |  |
| Unvaccinated | 1069 | 957 | 89.5 | 931 | 87.1 | 0.033 | 109 | 96 | 88.1 | 48 | 44.0 | 0.005 |
| 0-14 days of 1 dose | 635 | 559 | 88.0 | 543 | 85.5 |  | 33 | 26 | 78.8 | 15 | 45.5 |  |
| >14 days of 1 dose or 0-14 days of 2 doses | 269 | 223 | 82.9 | 218 | 81.0 |  | 587 | 465 | 79.2 | 310 | 52.8 |  |
| >14 days of 2 dose or 0-14d days of 3 doses | 56 | 37 | 66.1 | 36 | 64.3 |  | 511 | 402 | 78.7 | 265 | 51.9 |  |
| >14 days of 3 dose | - | - | - | - | - |  | 475 | 384 | 80.8 | 302 | 63.6 |  |
| Reinfection pre-vaccinated | - | - | - | - | - |  | 12 | 10 | 83.3 | 6 | 50.0 |  |
| ***p-value*** |  | 0.011 | | 0.034 | |  |  | 0.145 | | 0.341 | |  |

The significant was determined by Fisher extract’s test at *p*<0.05.

| **Table S5 Independent associations with symptom reporting between delta and omicron pandemic in aged>18 years** | | | | | | | | |
| --- | --- | --- | --- | --- | --- | --- | --- | --- |
| **Model** | **Jul-Oct 2021 (Delta) n=2029** | | | | **Jan-Mar 2022 (Omicron) n=1727** | | | |
|  | **Any symptoms** | | **Cough, fever, anosmia, ageusia** | | **Any symptoms** | | **Cough, fever, anosmia, ageusia** | |
|  | **OR (95% CI)** | ***p-value*** | **OR (95% CI)** | ***p-value*** | **OR (95% CI)** | ***p-value*** | **OR (95% CI)** | ***p-value*** |
| N gene (Ct) | 0.77  (0.56-0.86) | 0.000 | 0.90  (0.88-0.92) | 0.000 | 1.00  (0.98-1.02) | 0.915 | 0.95  (0.93-0.96) | 0.000 |
| ***Vaccination*** |  |  |  |  |  |  |  |  |
| Unvaccinated | ref. |  | ref. |  |  |  |  |  |
| 0-14 days of 1 dose | 0.79  (0.57-1.09) | 0.155 | 0.80  (0.60-1.08) | 0.153 | 0.34  (0.19-0.60) | 0.000 | 1.01  (0.46-2.24) | 0.979 |
| >14 days of 1 dose or 0-14 days of 2 doses | 0.51  (0.34-0.77) | 0.001 | 0.60  (0.41-0.89) | 0.012 | 0.65  (0.48-0.89) | 0.006 | 1.28  (0.83-1.96) | 0.265 |
| >14 days of 2 dose or 0-14 days of 3 doses | 0.25  (0.12-0.52) | 0.000 | 0.28  (0.13-0.58) | 0.001 | 0.58  (0.42-0.81) | 0.001 | 1.26  (0.82-1.95) | 0.287 |
| >14 days of 3 doses | **-** | **-** | **-** | **-** | 0.65  (0.46-0.92) | 0.014 | 2.14  (1.38-3.32) | 0.001 |
| Reinfection pre-vaccinated | **-** | **-** | **-** | **-** | 1.54  (0.35-6.78) | 0.566 | 1.70  (0.50-5.76) | 0.395 |

Data are represented as OR (95% CI), the significant was determined by binary logistic regression for age, gender, and comorbidity at *p*<0.05. OR, Odds ratio; CI, confidence interval; N gene, nucleocapsid gene; Ct, cycle threshold

| **Table S6 Independent factors of NCT of SARS-CoV-2 RNA by the multivariate Cox’s proportional hazard model by age >18 years (Jul-Oct 2021 (Delta))** | | | | |
| --- | --- | --- | --- | --- |
| **Factors** | **Survival analysis** | | | |
|  | **Adjusted Hazard Ratio**  **Exp(B)** | **95% CI** **for Exp(B)** | | ***p-value*** |
|  |  | **lower** **limit** | **detection limits** |  |
| Fever | 0.75 | 0.70 | 0.81 | <0.001* |
| Cough | 0.84 | 0.81 | 0.87 | <0.001* |
| Sore throat | 1.20 | 1.13 | 1.27 | 0.048* |
| Rhinorrhea | 1.20 | 1.14 | 1.26 | 0.027* |
| Productive sputum | 0.99 | 0.95 | 1.04 | 0.955 |
| Loss of taste | 0.95 | 0.89 | 1.01 | 0.921 |
| Loss of smell | 0.81 | 0.77 | 0.85 | <0001* |
| Dyspnea | 0.87 | 0.83 | 0.91 | 0.034* |
| Myalgia | 1.18 | 1.11 | 1.25 | <0.001* |
| Diarrhea | 1.37 | 1.24 | 1.52 | 0.503 |
| Nausea/vomiting | 1.09 | 0.96 | 1.24 | 0.154 |

Data are represented as Hazard Ratio (95% CI), the significant was determined by the Kaplan-Meier curves at *p*<0.05. NCT, negative conversion; SARS-CoV-2, severe acute respiratory syndrome coronavirus 2; CI, confidence interval

The Kaplan-Meier curves from multivariate Cox proportion estimating hazard rates between patients with and without symptom, adjusted age, gender and comorbidity were reported that patients who had systemic symptoms including fever (HR=0.75, 95% CI=0.70-0.80, *p*<0.001), cough (HR=0.84, 95% CI=0.81-0.87, *p*<0.001), and loss of smell (HR=0.81, 95% CI=0.77-0.85, *p*<0.001), had recovery rate greater than patients without these symptoms. On the other hand, patients with rhinorrhea (HR=1.20, 95% CI=1.14-1.26, *p*=0.027) or sore throat (HR=1.20, 95% CI=1.13-1.27, *p*=0.048) had recovery rate lower than another. The Kaplan-Meier curves revealed that fever, cough, and loss of smell had significantly prolonged NCT of SARS-CoV-2 RNA compared with the asymptomatic group (*p*<0.05).

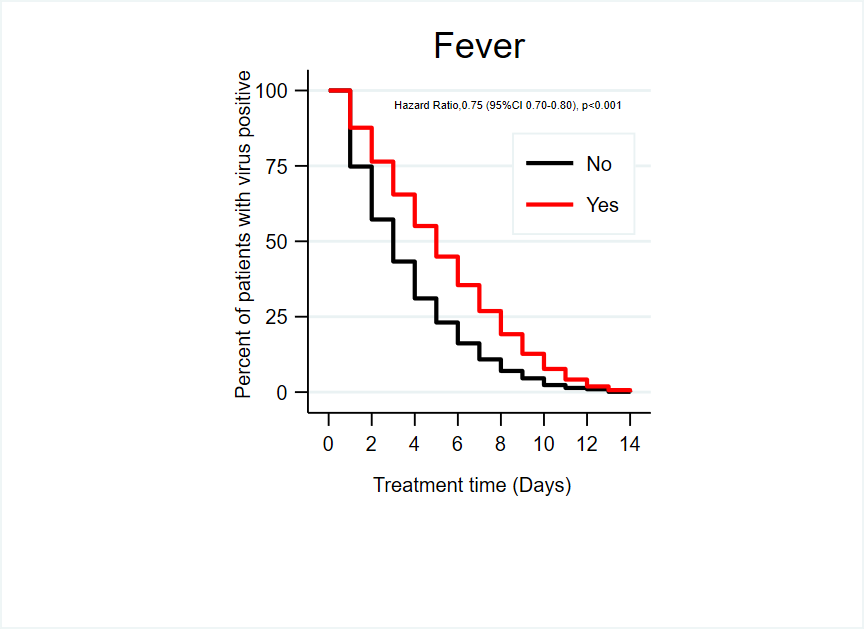

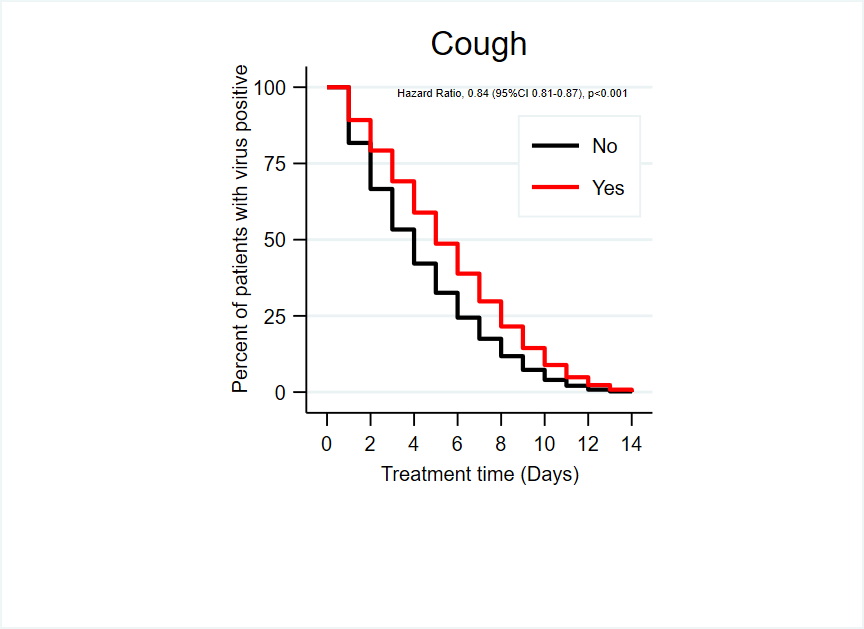

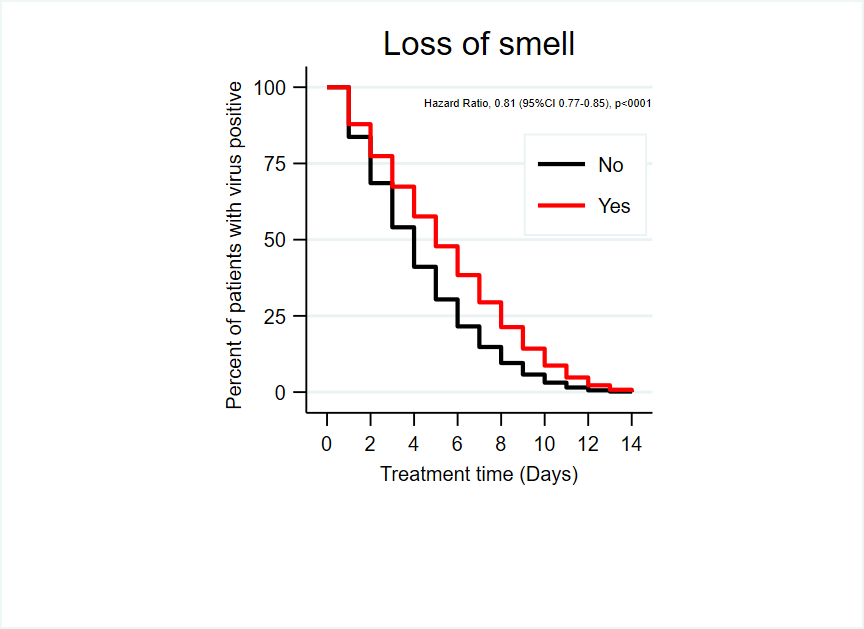

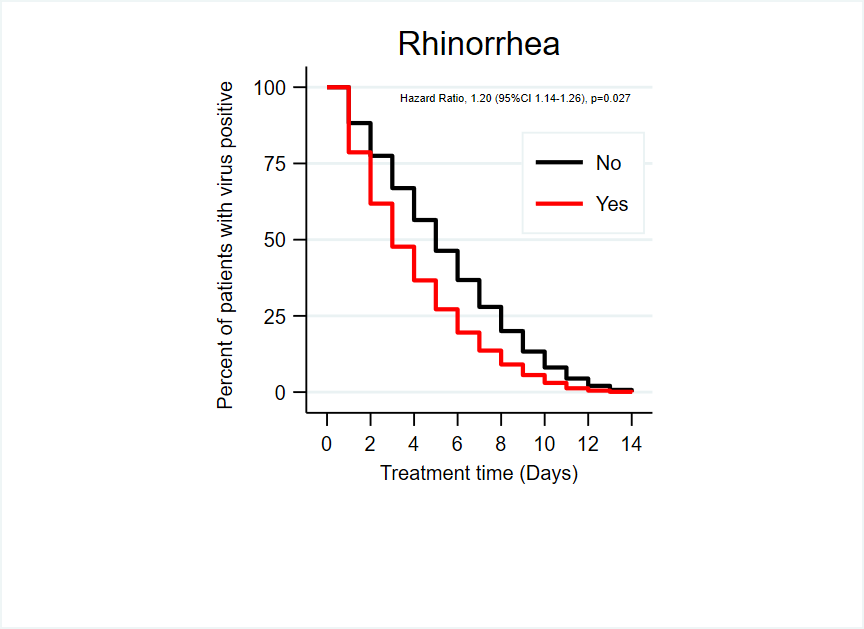

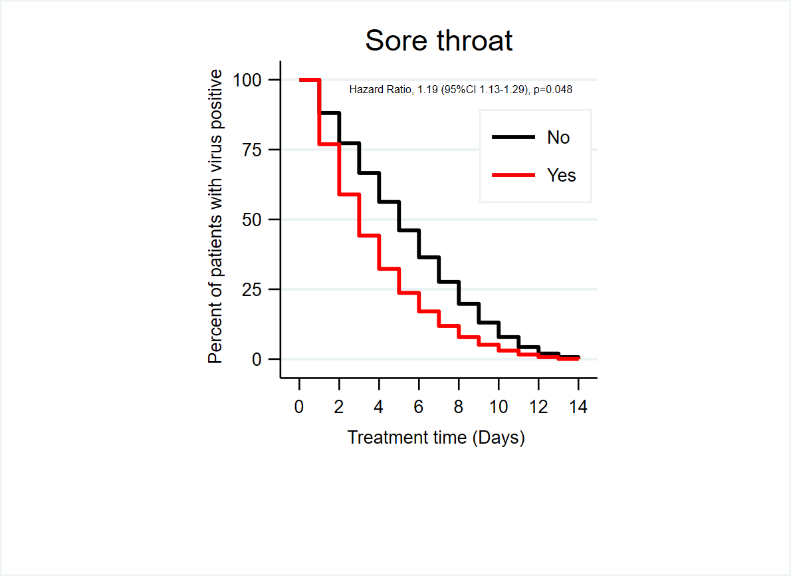

**Figure S3: Kaplan-Meier estimate of time to symptom alleviation as reported by Covid-19-adapted FLU-PRO questionnaire (Jul-Oct 2021 (Delta))**

Kaplan-Meier Estimate for treatment time (days) and percentage of patients infected with Delta in July-October 2022 by using self-reported symptoms questionnaire: patients had the symptom in red line and patients did not have symptom in black line.

| **Table S7 Characteristics of the participants and geometric mean humoral immunogenicity assay responses according to anti-SARS-CoV-2 nucleocapsid protein and vaccine group** | | | | | | | | | | | | | | | | | |
| --- | --- | --- | --- | --- | --- | --- | --- | --- | --- | --- | --- | --- | --- | --- | --- | --- | --- |
| **Characteristics** | | **Anti-SARS-CoV-2 NP** | | | | | | | **Vaccine Group** | | | | | | | | |
|  |  | **Total**  **(n=337)** | | **Negative**  **(n=116)** | | **Positive**  **(n=221)** | | ***p-value*** | **Total**  **(n=495)** | | **Unvaccinated**  **(n=213)** | | **ChAdOx1**  **(n=182)** | | **CoronaVac**  **(n=100)** | | ***p-value*** |
|  |  | n | % | n | % | n | % |  | n | % | n | % | n | % | n | % |  |
| Gender | |  |  |  |  |  |  |  |  |  |  |  |  |  |  |  |  |
| Male | | 123 | 36.5 | 43 | 37.1 | 80 | 36.2 | 0.483 | 179 | 36.2 | 81 | 38.0 | 64 | 35.2 | 34 | 34.0 | 0.723 |
| Female | | 214 | 63.5 | 73 | 62.9 | 141 | 63.8 |  | 316 | 63.8 | 132 | 62.0 | 118 | 64.8 | 66 | 66.0 |  |
| Age, years, median (range) | | 43 | (31-55) | 39 | (28-50.5) | 44 | (33-56) | 0.012 | 43 | (30-56) | 40 | (24-53) | 49 | (36-61) | 39 | (30-48) | <0.001 |
|  | 12-18 | 24 | 7.1 | 14 | 12.1 | 10 | 4.5 | 0.029 | 37 | 7.5 | 35 | 16.4 | 0 | 0.0 | 1 | 1.0 | <0.001 |
|  | 19-45 | 167 | 49.6 | 58 | 50.0 | 109 | 49.3 |  | 238 | 48.1 | 94 | 44.1 | 79 | 43.7 | 65 | 65.0 |  |
|  | >45 | 146 | 43.3 | 44 | 37.9 | 102 | 46.2 |  | 220 | 44.4 | 84 | 39.4 | 102 | 56.4 | 34 | 34.0 |  |
| ***Presence of comorbidities*** | | |  |  |  |  |  |  |  |  |  |  |  |  |  |  |  |
|  | Hypertension | 49 | 14.5 | 17 | 14.7 | 32 | 14.5 | 0.559 | 78 | 15.8 | 30 | 14.1 | 37 | 20.3 | 11 | 11.0 | 0.064 |
|  | Dyslipidemia | 26 | 7.7 | 11 | 9.5 | 15 | 6.8 | 0.265 | 38 | 7.7 | 7 | 3.3 | 23 | 12.6 | 8 | 8.0 | 0.001 |
|  | Diabetes mellitus | 25 | 7.4 | 7 | 6.0 | 18 | 8.1 | 0.304 | 38 | 7.7 | 15 | 7.0 | 19 | 10.4 | 4 | 4.0 | 0.136 |
|  | Obesity | 5 | 1.5 | 1 | 0.9 | 4 | 1.8 | 0.430 | 4 | 0.8 | 1 | 0.5 | 2 | 1.1 | 1 | 1.0 | 0.682 |
|  | Asthma/COPD | 18 | 5.3 | 4 | 3.5 | 14 | 6.3 | 0.441 | 28 | 5.7 | 10 | 4.7 | 13 | 7.1 | 5 | 5.0 | 0.769 |
|  | Chronic heart disease | 8 | 2.4 | 1 | 0.9 | 7 | 3.2 | 0.208 | 9 | 1.8 | 4 | 1.9 | 4 | 2.2 | 1 | 1.0 | 0.830 |
|  | Others | 67 | 19.9 | 24 | 20.7 | 43 | 19.5 | 0.446 | 93 | 18.8 | 49 | 23.0 | 33 | 18.1 | 11 | 11.0 | 0.037 |
| ***Vaccine manufacturer*** | | |  |  |  |  |  |  |  |  |  |  |  |  |  |  |  |
|  | Unvaccinated | 139 | 41.2 | 51 | 44.0 | 88 | 39.8 | 0.005 | - | - | - | - | - | - | - | - | na |
|  | ChAdOx1 | 126 | 37.4 | 52 | 44.8 | 74 | 33.5 |  | - | - | - | - | - | - | - | - |  |
|  | CoronaVac/ BBIBP-CorV | 72 | 21.4 | 13 | 11.2 | 59 | 26.7 |  | - | - | - | - | - | - | - | - |  |
| ***Vaccine doses administered*** | | |  |  |  |  |  |  |  |  |  |  |  |  |  |  |  |
|  | Unvaccinated | 139 | 41.2 | 51 | 44.0 | 88 | 39.8 | <0.001 | 213 | 43.0 | 213 | 100.0 | 0 | 0.0 | 0 | 0.0 | <0.001 |
|  | 1 dose | 102 | 30.3 | 48 | 41.4 | 54 | 24.4 |  | 158 | 31.9 | 0 | 0.0 | 143 | 78.6 | 15 | 15.0 |  |
|  | 2 doses | 76 | 22.6 | 15 | 12.9 | 61 | 27.6 |  | 100 | 20.2 | 0 | 0.0 | 15 | 8.2 | 85 | 85.0 |  |
|  | 3 doses | 20 | 5.9 | 2 | 1.7 | 18 | 8.1 |  | 24 | 4.8 | 0 | 0.0 | 24 | 13.2 | 0 | 0.0 |  |
| ***SARS-CoV-2*** | |  |  |  |  |  |  |  |  |  |  |  |  |  |  |  |  |
|  | seronegative | - | - | - | - | - | - | na | 116 | 34.4 | 51 | 36.7 | 52 | 41.3 | 13 | 18.1 | 0.002 |
|  | seropositive | - | - | - | - | - | - |  | 221 | 65.6 | 88 | 63.3 | 74 | 58.7 | 59 | 81.9 |  |
| Immunoglobulin G (mg/ml); GMT, (95%CI) | | 809.8 | (696-  942.3) | 1079 | (918.6-1268) | 470.9 | (351.2-631.5) | <0.001 | 797.3 | (702.9-904.4) | 587 | (463.9-742.7) | 1094 | (950.7-1260) | 884.9 | (701-1117) | 0.002 |
| Neutralizing antibody; GMT (95%CI) | | 97.5 | (94.7-  98.2) | 70.7 | (61.3-81.6) | 91.6 | (89.1-94.2) | <0.001 | 97.4 | (94.6-98.0) | 78.9 | (74.4-83.6) | 94.8 | (92.2-97.5) | 93.3 | (90.2-96.4) | <0.001 |
| Days since most recent positive viral test, median (range) | | 89 | (71.5-  120) | 102 | (79-124) | 84 | (69-114) | 0.007 | 98 | (81-107) | 99 | (94-120) | 91 | (70-100) | 98 | (84-  107) | <0.001 |
| Days since receipt of first vaccine dose, median (range) | | 135 | (114-  159) | 135 | (119-162) | 135 | (113-159) | 0.884 | 133 | (117-151) | - | - | 133 | (117-150) | 128 | (114-198) | 0.076 |
| Days since receipt of second vaccine dose, median (range) | | 132 | (63-  177) | 59 | (34.5-171) | 142 | (92-181) | 0.004 | 136 | (65.5-179) | - | - | 55 | (33-117) | 170 | (136-187) | <0.001 |
| Days since receipt of third vaccine dose, median (range) | | 109 | (88-125) | 103 | (102-104) | 111 | (88-125) | 0.739 | 118 | (109-137) | - | - | 118 | (109 - 137) | - | - | na |
| ***Cycle Threshold (viral load at the time of entering HI)*** | | | | | | | | | | | | | | | | | |
| Nucleocapsid (N), median (range) | | 20.0 | (17.2-23.8) | 20.9 | (17.9-26.5) | 19.2 | (16.8-22.9) | 0.049 | 20.2 | (17.5-24.6) | 20.4 | (17.5-25.8) | 20.1 | (17.4-24.2) | 20.0 | (16.8-24.6) |  |
|  | <20 | 153 | 50.5 | 45 | 42.1 | 108 | 55.1 | 0.004 | 212 | 48.0 | 93 | 45.8 | 81 | 49.7 | 38 | 50.0 | 0.587 |
|  | 20-30 | 125 | 41.3 | 46 | 43.0 | 79 | 40.3 |  | 192 | 43.4 | 95 | 46.8 | 68 | 41.7 | 29 | 38.2 |  |
|  | >30 | 25 | 8.3 | 16 | 15.0 | 9 | 4.6 |  | 38 | 8.6 | 15 | 7.4 | 14 | 8.6 | 9 | 11.8 |  |
| Envelope (E), median (range) | | 16.8 | (13.4-21.1) | 17.7 | (14.0-23.0) | 16.3 | (13.2-19.9) | 0.040 | 17.0 | (13.6-21.6) | 17.2 | (13.5-22.2) | 16.5 | (13.5-21.1) | 17.1 | (14.2-22.3) |  |
|  | <20 | 231 | 71.3 | 71 | 63.4 | 160 | 75.5 | 0.028 | 325 | 69.3 | 142 | 69.3 | 116 | 69.5 | 67 | 69.1 | 0.286 |
|  | 20 - 30 | 80 | 24.7 | 33 | 29.5 | 47 | 22.2 |  | 128 | 27.3 | 58 | 28.3 | 47 | 28.1 | 23 | 23.7 |  |
|  | >30 | 13 | 4.0 | 8 | 7.1 | 5 | 2.4 |  | 16 | 3.4 | 5 | 2.4 | 4 | 2.4 | 7 | 7.2 |  |
| RNA-dependent RNA polymerase (RdRp), median (range) | | 21.5 | (18.4- 25.1) | 21.8 | (19.1 -28.2) | 20.9 | (18.0- 24.2) | 0.004 | 21.7 | (18.7- 26.1) | 21.9 | (18.7- 27.2) | 21.2 | (18.6- 25.4) | 21.6 | (18.8-26.1) | 0.778 |

Categorical and continuous data of characteristics of the participants and geometric mean humoral immunogenicity assay responses presented as n (%) and median (range)at *p*<0.05 indicates statistical significance. COPD, chronic obstructive pulmonary disease; GMC, geometric mean concentration; ChAdOx1, AstraZeneca vaccine; CoronaVac, Sinovac vaccine; BBIBP-CorV, Sinopharm vaccine; SARS-CoV-2, the severe acute respiratory syndrome-coronavirus-2; NP, nucleocapsid protein; CI, confident intervals; Ct, cycle threshold; na, not available.

| **Table S8 Anti-RBD IgG and sVNT geometric mean concentration (GMC) among unvaccinated, ChAdOx1, and CoronaVac** | | | | | | | | | | | | | | | | | | | | |
| --- | --- | --- | --- | --- | --- | --- | --- | --- | --- | --- | --- | --- | --- | --- | --- | --- | --- | --- | --- | --- |
| **Vaccinations/Day** | | **Unvaccinated** | | | **ChAdOx1** | | | | | | | | | **CoronaVac** | | | | | | ***p-value*** |
|  |  | **25-56 days** | **56-84 days** | **>84 days** | **25-56 days** | | | **56-84 days** | | | **>84 days** | | | **25-56 days** | | **56-84 days** | | **>84 days** | |  |
|  |  |  |  |  | **1 dose** | **2 doses** | **3 doses** | **1 dose** | **2 doses** | **3 doses** | **1 dose** | **2 doses** | **3 doses** | **1 dose** | **2 doses** | **1 dose** | **2 doses** | **1 dose** | **2 doses** |  |
| Number of objectives | | 22 | 42 | 149 | 19 | 10 | 10 | 40 | 12 | 11 | 49 | 18 | 13 | 9 | 10 | 12 | 14 | 16 | 39 |  |
| ***SARS-CoV-2 IgG*** | |  | | | | | | | | | | | | | | | | | | 0.022 |
|  | GMT of IgG | 299.5 | 597.2 | 645.1 | 1655 | 1440 | 1538 | 1025 | 2075 | 1674 | 822.9 | 945.3 | 886.1 | 566.8 | 897.6 | 789.1 | 712.1 | 1174 | 974.5 |  |
|  | Lower 95% CI | 140.9 | 339.6 | 488.4 | 1217 | 889 | 861.7 | 742.6 | 1057 | 1147 | 626.4 | 567.4 | 588.2 | 252 | 450.6 | 305.5 | 315.3 | 638.4 | 684.9 |  |
|  | Upper 95% CI | 636.6 | 1050 | 852 | 2251 | 2332 | 2747 | 1414 | 4074 | 2443 | 1081 | 1575 | 1335 | 1275 | 1788 | 2038 | 1608 | 2158 | 1386 |  |
|  | ***p-value*** | 0.044 | | | *ns* | | | *ns* | | | *ns* | | | *ns* | | *ns* | | *ns* | |  |
| ***SARS-CoV-2 sVNT*** | |  | | | | | | | | | | | | | | | | | | <0.0001 |
|  | GMT of sVNT | 74.3 | 75.7 | 80.5 | 97.8 | 97.4 | 97.3 | 90.2 | 97.4 | 98.0 | 95.2 | 95.1 | 96.4 | 92.7 | 92.8 | 87.1 | 86.8 | 95.6 | 97.2 |  |
|  | Lower 95% CI | 62.5 | 63.3 | 75.6 | 97.4 | 96.6 | 96.4 | 79.8 | 96.6 | 97.7 | 93.9 | 93.0 | 94.8 | 87.3 | 86.5 | 73.7 | 71.1 | 92.6 | 96.6 |  |
|  | Upper 95% CI | 88.3 | 90.6 | 85.6 | 98.2 | 98.3 | 98.2 | 102.0 | 98.2 | 98.3 | 96.6 | 97.2 | 97.9 | 98.5 | 99.4 | 103.0 | 105.9 | 98.8 | 97.7 |  |
|  | ***p-value*** | *ns* | | | *ns* | | | *ns* | | | *ns* | | | *ns* | | *ns* | | *ns* | |  |

Relation of vaccinations ChAdOx1 (AstraZeneca), CoronaVac (Sinovac), Unvaccinated and laboratory findings of all confirmed COVID-19 patients. Presented as vaccinated 25-56 days, 56-84 days, >84 days and range at *p*<0.05 indicates statistical significance. SARS-CoV-2 IgG quantitative immunological test. By examining the ability to prevent the severity of symptoms when infected. SARS-CoV-2 sVNT Immunity test by examining the ability to inhibit invasion and viral proliferation using the Surrogate virus Neutralization test (sVNT). SARS_CoV-2, the severe acute respiratory syndrome-Coronavirus-2; IgG, immunoglobulin G; GMT, geometric mean titer; sVNT, the surrogate virus neutralization test

| **Table S9 Anti-RBD IgG and sVNT geometric mean concentration (GMC) among unvaccinated, ChAdOx1, and CoronaVac by age** | | | | | | | | |
| --- | --- | --- | --- | --- | --- | --- | --- | --- |
| **Vaccinations/Age** | | **Unvaccinated** | | **ChAdOx1** | | **CoronaVac** | | ***p-value*** |
|  |  | **Age<45** | **Age≥45** | **Age<45** | **Age≥45** | **Age<45** | **Age≥45** |  |
| Number of objectives | | 129 | 84 | 80 | 102 | 66 | 34 |  |
| ***SARS-CoV-2 IgG*** | |  |  |  |  |  |  | <0.001 |
|  | GMT of IgG | 486 | 784.2 | 830.8 | 1418 | 822.4 | 1020 |  |
|  | Lower 95% CI of GMT | 350.1 | 570.5 | 682.4 | 1188 | 608.8 | 700.8 |  |
|  | Upper 95% CI of GMT | 674.6 | 1078 | 1012 | 1692 | 1111 | 1484 |  |
|  | ***p-value*** | 0.001 | | 0.004 | | *ns* | |  |
| ***SARS-CoV-2 sVNT*** | |  |  |  |  |  |  | <0.001 |
|  | GMT of sVNT | 74.5 | 85.9 | 92.5 | 96.9 | 92.3 | 95.2 |  |
|  | Lower 95% CI of GMT | 68.3 | 81.2 | 87.1 | 96.4 | 87.8 | 93.3 |  |
|  | Upper 95% CI of GMT | 81.4 | 90.9 | 98.3 | 97.5 | 97.0 | 97.0 |  |
|  | ***p-value*** | 0.034 | | *ns* | | *ns* | |  |

Relation of vaccination ChAdOx1 (AstraZeneca), CoronaVac (Sinovac), Unvaccinated and laboratory findings of all confirmed COVID-19 patients. Presented patients age <45 and ≥45 years range at *p*<0.05 indicates statistical significance. SARS-CoV-2 IgG quantitative immunological test. By examining the ability to prevent the severity of symptoms when infected. SARS-CoV-2 sVNT Immunity test by examining the ability to inhibit invasion and viral proliferation using the Surrogate virus Neutralization test (sVNT). Anti-RBD IgG, anti-receptor binding domain immunoglobulin G; IgG, immunuglobulin G; SARS-CoV-2, the severe acute respiratory syndrome-coronavirus-2; sVNT, the surrogate virus neutralization test; GMT, Geometric mean titer; ns, not statistically significant.

| **Table S10 SARS-CoV-2 sVNT geometric mean concentration (GMC) among unvaccinated, ChAdOx1, and CoronaVac by Wuhan and Delta variants** | | | | | | | | | | | | | | |
| --- | --- | --- | --- | --- | --- | --- | --- | --- | --- | --- | --- | --- | --- | --- |
| **Days/Age** | | **25-56 days** | | | | **56-84 days** | | | | **>84 days** | | | | ***p-value*** |
|  |  | **Age <45** | | **Age ≥45** | | **Age <45** | | **Age ≥45** | | **Age <45** | | **Age ≥45** | |  |
|  |  | **Wuhan** | **Delta** | **Wuhan** | **Delta** | **Wuhan** | **Delta** | **Wuhan** | **Delta** | **Wuhan** | **Delta** | **Wuhan** | **Delta** |  |
| **Unvaccinated** | |  |  |  |  |  |  |  |  |  |  |  |  | <0.0001 |
|  | Number of subjects | 12 | 9 | 10 | 8 | 27 | 15 | 26 | 15 | 88 | 60 | 84 | 59 |  |
|  | GMT of sVNT | 72.1 | 77.4 | 18.9 | 32.2 | 69.5 | 88.3 | 29.0 | 47.6 | 76.5 | 86.6 | 37.0 | 48.0 |  |
|  | Lower 95% CI of GMT | 55.2 | 59.8 | 3.2 | 9.6 | 53.1 | 76.5 | 15.3 | 27.3 | 69.6 | 81.2 | 26.8 | 34.8 |  |
|  | Upper 95% CI of GMT | 94.1 | 100.2 | 113.0 | 108.3 | 91.0 | 101.9 | 55.0 | 83.1 | 84.1 | 92.4 | 51.1 | 66.1 |  |
| ***p-value*** | | 0.021 | | 0.034 | | *ns* | | *ns* | | 0.015 | | 0.004 | |  |
| **ChAdOx1** | |  |  |  |  |  |  |  |  |  |  |  |  | <0.0001 |
|  | Number of subjects | 15 | 24 | 15 | 24 | 30 | 33 | 29 | 33 | 35 | 45 | 35 | 45 |  |
|  | GMT of sVNT | 97.2 | 97.8 | 75.4 | 89.1 | 88.0 | 97.5 | 63.8 | 79.7 | 94.6 | 96.0 | 47.9 | 71.1 |  |
|  | Lower 95% CI of GMT | 96.4 | 97.5 | 61.7 | 84.0 | 74.6 | 97.1 | 47.3 | 69.8 | 93.0 | 94.9 | 35.7 | 59.3 |  |
|  | Upper 95% CI of GMT | 97.9 | 98.2 | 92.2 | 94.6 | 103.8 | 97.9 | 86.1 | 91.0 | 96.2 | 97.2 | 64.3 | 85.2 |  |
| ***p-value*** | | 0.032 | | ns | | 0.028 | | 0.007 | | 0.0001 | | 0.001 | |  |
| **CoronaVac** | |  |  |  |  |  |  |  |  |  |  |  |  | <0.0001 |
|  | Number of subjects | 12 | 11 | 12 | 11 | 18 | 8 | 18 | 8 | 35 | 15 | 35 | 15 |  |
|  | GMT of sVNT | 84.6 | 94.2 | 24.3 | 63.7 | 94.6 | 94.4 | 63.5 | 56.3 | 94.0 | 96.3 | 71.2 | 68.6 |  |
|  | Lower 95% CI of GMT | 66.8 | 90.5 | 8.6 | 42.3 | 91.1 | 88.0 | 43.1 | 24.4 | 89.0 | 94.6 | 54.0 | 51.7 |  |
|  | Upper 95% CI of GMT | 107.1 | 98.1 | 68.6 | 95.9 | 98.2 | 101.2 | 93.4 | 129.9 | 99.3 | 97.9 | 93.7 | 91.2 |  |
| ***p-value*** | | 0.001 | | 0.013 | | 0.038 | | 0.009 | | 0.007 | | 0.027 | |  |

Represents the mean sVNT geometric mean concentration (GMC) of unvaccinated, AstraZeneca COVID-19 (ChAdOx1) vaccinated, and Sinovac COVID-19 (CoronaVac) vaccinated patients. Comparing each vaccination group and each age group. During the Wuhan and Delta outbreaks, there was no difference in sVNT values. In addition to the AstraZeneca COVID-19 (ChAdOx1) vaccinated before infection >84 days in the age range ≥45 years, were found statistical differences. SARS-CoV-2, the severe acute respiratory syndrome-coronavirus-2; sVNT, the surrogate virus neutralization test; GMC, geometric mean concentration; CI, confidence interval; ns, not statistically significant.

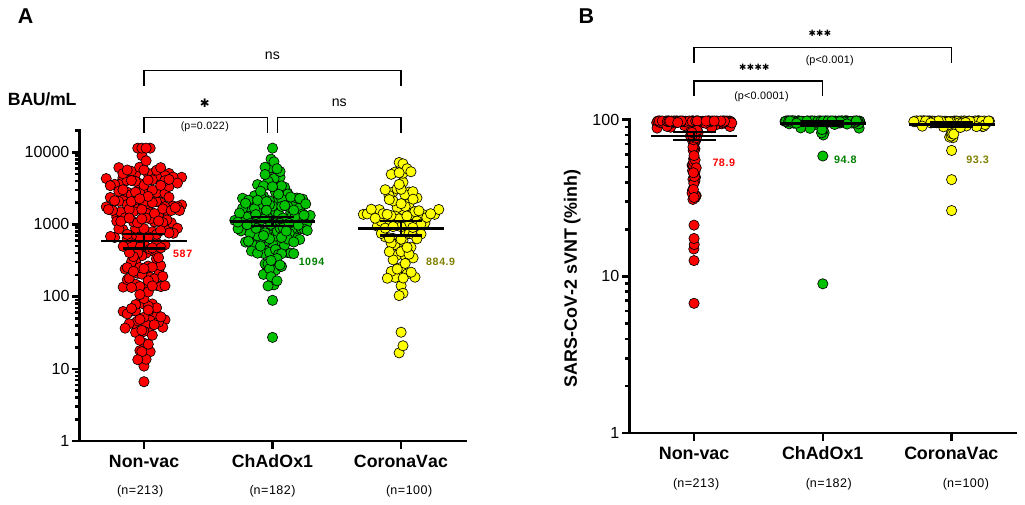

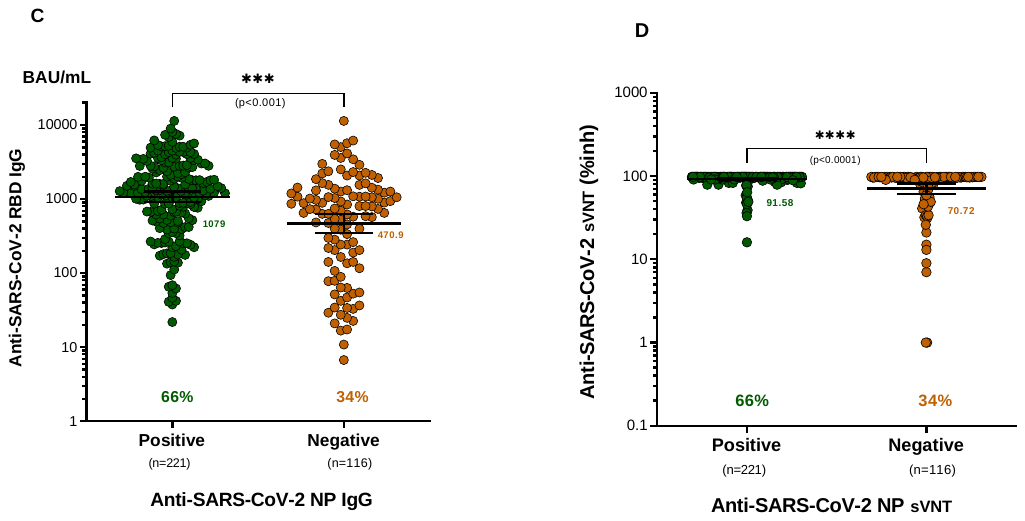

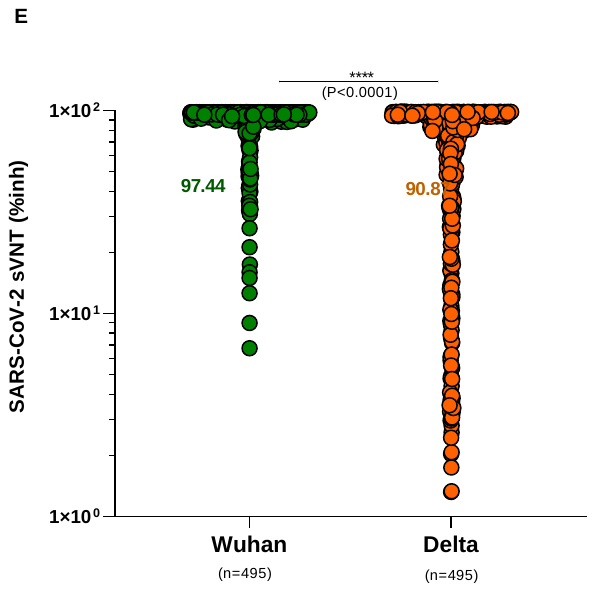

**Figure S4-Plasma Anti-Immunoglobulin G (IgG) and Antibody (sVNT) levels against SARS**

A	Anti-SARS-CoV-2 spike receptor binding domain (RBD) IgG antibodies in plasma of unvaccinated cases (n=213), ChAdOx1 nCoV-19 vaccinated cases (n=182), and CoronaVac vaccinated cases (n=100) were measured by chemiluminescent microparticle immunoassay.

B	Anti-SARS-CoV-2 neutralizing antibodies in plasma of unvaccinated cases (n=213), ChAdOx1 nCoV-19 vaccinated cases (n=182), and CoronaVac vaccinated cases (n=100) were measured by surrogate virus neutralization test (sVNT).

C	Anti-SARS-CoV-2 spike RBD IgG antibodies in plasma of positive anti-SARS-CoV-2 nucleocapsid protein cases (n=221) and negative anti-SARS-CoV-2 nucleocapsid protein cases (n=116) were determined by chemiluminescent microparticle immunoassay and SARS-CoV-2 IgG II Quant for use with ARCHITECT; Abbott Laboratories.

D	Anti-SARS-CoV-2 neutralizing antibodies in plasma of positive anti-SARS-CoV-2 nucleocapsid protein cases (n=221) and negative anti-SARS-CoV-2 nucleocapsid protein cases (n=116) were measured by surrogate virus neutralization test (sVNT) and SARS-CoV-2 IgG II Quant for use with ARCHITECT; Abbott Laboratories.

E	Anti-SARS-CoV-2 neutralizing antibodies in plasma of wild-type (Wuhan) strain infection cases (n=495) and Delta (B.1.617.2) strain infection cases (n=495) were measured by surrogate virus neutralization test (sVNT).

Data information

A-B: Statistical comparisons were determined by two-tailed Kruskal-wallis test.

C-E: Statistical comparisons were determined by two-tailed Mann-Whitney U-test.

In all graphs, error bars denote median and interquartile range.
